# An Environmental Vulnerability Index framework supporting targeted public health interventions at the census tracts level

**DOI:** 10.1101/2024.10.16.24315575

**Authors:** Lauren B. Anderson, Rochelle H. Holm, Caison Black, Donald J. Biddle, Weihsueh A. Chiu, Aruni Bhatnagar, Ted Smith

## Abstract

**BACKGROUND:** Analyzing and visualizing disparities in environmental risks can help in assessing place-based vulnerabilities and provide civic leaders and community members with essential data about promoting health equity and inform public health strategies. However, there is a lack of effective and integrative tools for evaluating census tract vulnerabilities.

**OBJECTIVE:** We investigated the adaption of a previously developed environmental vulnerability index to evaluate cumulative impacts of diverse stressors in Louisville Metro-Jefferson County, KY, with the goal of supporting multi-faceted targeted public health interventions at the census tract-level.

**METHODS:** We assessed countywide variability in vulnerability using Toxicological Prioritization Index interface across five domains with 32 indicators and modeled the effects of theoretical public health interventions.

**RESULTS:** Our findings suggest similarly vulnerable areas are not always geographically clustered. Higher vulnerability scores are observed along the western and central areas of the county with lower vulnerability scores in the central urban core and eastern regions. The index enabled the selection of the most at-risk census tracts for modeling targeted public health interventions to reduce cumulative environmental vulnerability.

**SIGNIFICANCE:** Environmental vulnerabilities are not invariant features of urban environments, rather the knowledge of these risks can guide the development and implementation of targeted solutions.

**IMPACT STATEMENT:** Targeted interventions to modify environmental conditions that are supportive of health can be developed and implemented locally with greater precision at the census tract level, yielding impactful outcomes.

## Introduction

Given the importance of the role that environmental factors play in health outcomes, assessing and mapping population health and environmental hazards together could better estimate place-based vulnerabilities and furnish civic leaders and community members with vital data on health equity and environmental risks. This knowledge can facilitate informed decisions regarding the implementation of public health interventions tailored to address specific vulnerabilities within communities. However, addressing the unequal distribution of environmental hazards and vulnerabilities across geographic areas presents multifaceted challenges.^1–4^ Health risk and environmental hazards are well described at the state and county levels, but the characteristics that comprise vulnerability are often localized at the census tract or neighborhood level.^5^ For instance, infectious disease tracking by the National Notifiable Diseases Surveillance System^6^ happens at the state level despite the fact that individual disease occurrence is shaped by geographical proximity at a city or a neighborhood scale.^7^ Similarly, some determinants of health, such as access to healthy foods, or proximity to only fast food and convenience outlets, are well established to be most impactful at a neighborhood scale.^8^ Further, risk and vulnerability do not recognize political boundaries; pollution can cross census tract, neighborhood, and even state boundaries.^9^ However, there is paucity of integrative, census tract, vulnerability screening tools to inform targeted public health interventions.

Existing vulnerability indices at smaller scales typically focus on a relatively narrow set of issues such as environmental pollution, extreme heat, flooding, disease, or lead exposure and depend on specific data sources which may limit appropriate use and effectiveness.^10–14^ The World Health Organization (WHO) provides a framework for global application to evaluate the cost-effectiveness of environmental health interventions for air pollution, water supply, sanitation, climate change, food safety, water management, and vector control;^15^ while lacking place-based intervention framework customization. Moreover, literature regarding public health interventions to address climate-related environmental vulnerabilities and extreme weather events is lacking.^16^ Many of these evaluations focus on modifying individual stressors or exposures to individuals, rather than using a place-based framework that integrates cumulative impacts, community exposure, and the natural and built environments.

As health inequities continue to grow nationally and new vulnerabilities arise from a changing climate, frameworks that integrate environmental hazard and risk data to understand vulnerability will become increasingly important. There are some efforts to integrate national and local health data in a spatial context to address localized concerns across several environmental domains.^1^ For instance, the Houston–Galveston–Brazoria (HGB) EnviroScreen’s Environmental Vulnerability Index (EVI)^1^ pinpoints which communities need the most support by analyzing health and environmental data geographically. This index preceded the U.S. Climate Vulnerability Index, which integrates indictors nationwide to inform a broad range of policy interventions ranging from health and environment to infrastructure and socio-economic factors.^2,17^ A potential benefit of environmental vulnerability assessments includes the prioritization and implementation of layered interventions to reduce cumulative burdens across communities. By employing comprehensive screening tools that merge publicly accessible health data with location-specific environmental indicators, vulnerabilities can be pinpointed. This approach forms a strong foundation for implementing precise public health intervention strategies. The goal of this study was to demonstrate the development of an integrative screening tool to inform targeted public health interventions at a metro-area scale. Specifically, we adapted the HGBEnviroScreen framework into a bespoke index for Louisville Metro-Jefferson County. We used the framework to model and evaluate interventions to reduce environmental risk and vulnerabilities, focusing on solutions for communities most burdened by multiple stressors.

## Materials and Methods

### Study Area

The Louisville Metro-Jefferson County area in Kentucky, USA, is a mid-sized, metropolitan area with a population of 780,000.^18^ Employment is largely in trade, transportation, and utilities.^19^ Manufacturing activities are dispersed throughout the county, including a chemical and rubber manufacturing corridor along the western edge of the city. The county features mixed-income housing in the north and west, high-income areas in the east, and middle-to-low-income zones in the south. Many census tracts in the northwestern part of the county are identified by the Climate and Economic Justice Screening Tool^20^ as facing significant burdens. The people, environment, and infrastructure covering 190 census tracts are additionally affected by the presence of federal and state Superfund sites, an international airport with a commercial air-transport hub, two large interstate highways, and the Ohio River abutting the northern and western boundaries of the county (Figure 1).

**Figure 1.**
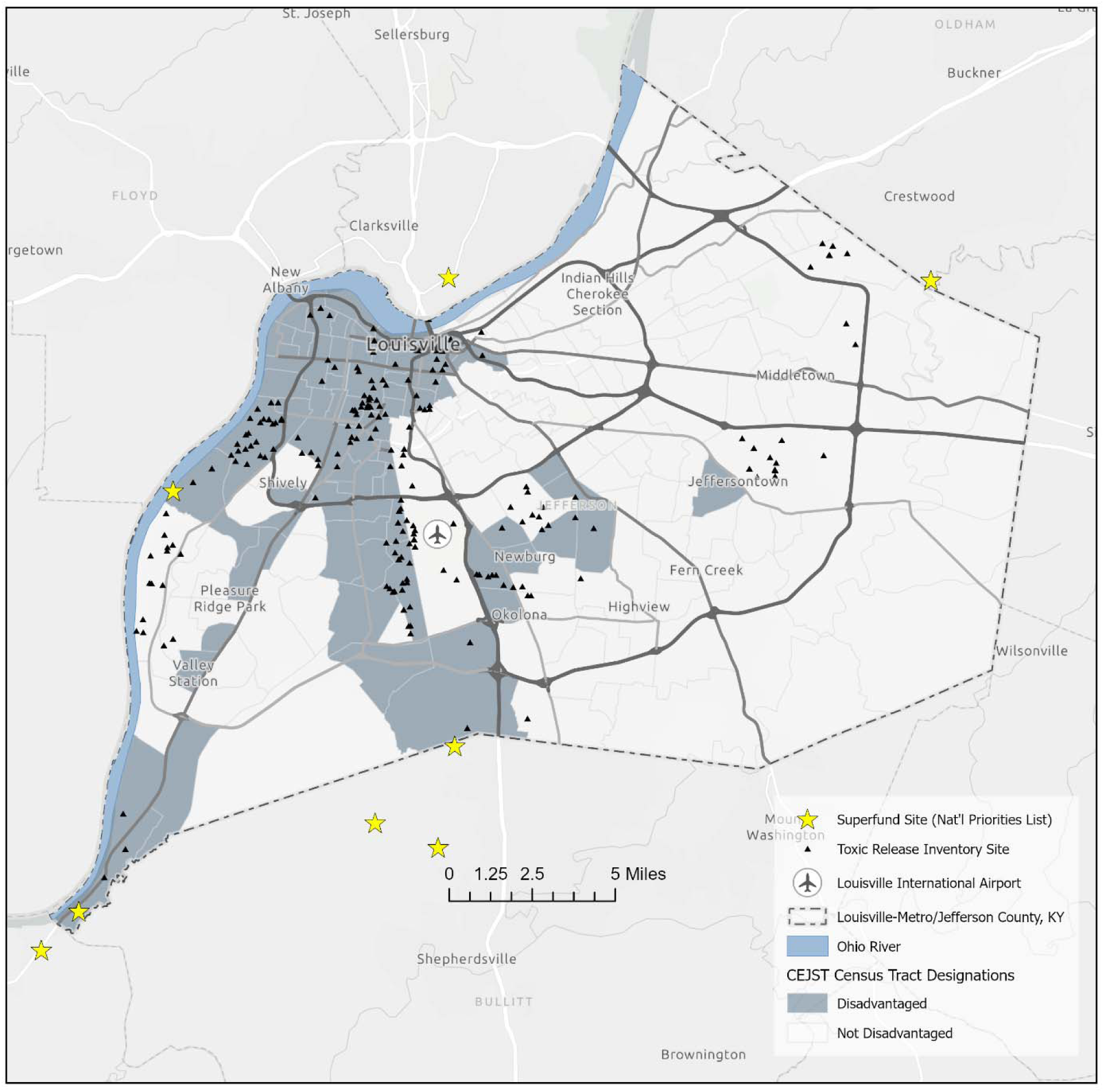
Study area, Louisville Metro-Jefferson County area in Kentucky, USA. The Climate and Economic Justice Screening Tool (CEJST) designation by census tract as well as federal and state Superfund sites, the international airport with a commercial air-transport hub, two large interstate highways, and the Ohio River on the north are represented.

### Data Sources

Our methodological approach was based on the HGB region tool^1^ (Table S1) with customization to reflect Louisville Metro-Jefferson County-relevant indicators. The index includes 32 indicators organized into five domains (Table 1; Table S2). Transportation noise exposure (vehicle, railways, aviation), heat wave exposure, and tornado indicators were added. Some indicators included in the original HGB index were removed for the Jefferson County index as being either not applicable or with no local source data (Table S3). Indicators were derived from source datasets that were publicly available, with source data from 2015 through 2023. For proximity analysis including number of hospitals, Superfund sites, and point sources of pollution, a sum of sites within a 5 km radius buffer from the census tract centroid was used. In most cases, low indicator values reflect low vulnerability. This relationship was inverted for three indicators (life expectancy, number of hospitals, and tree canopy).

**Table 1.**
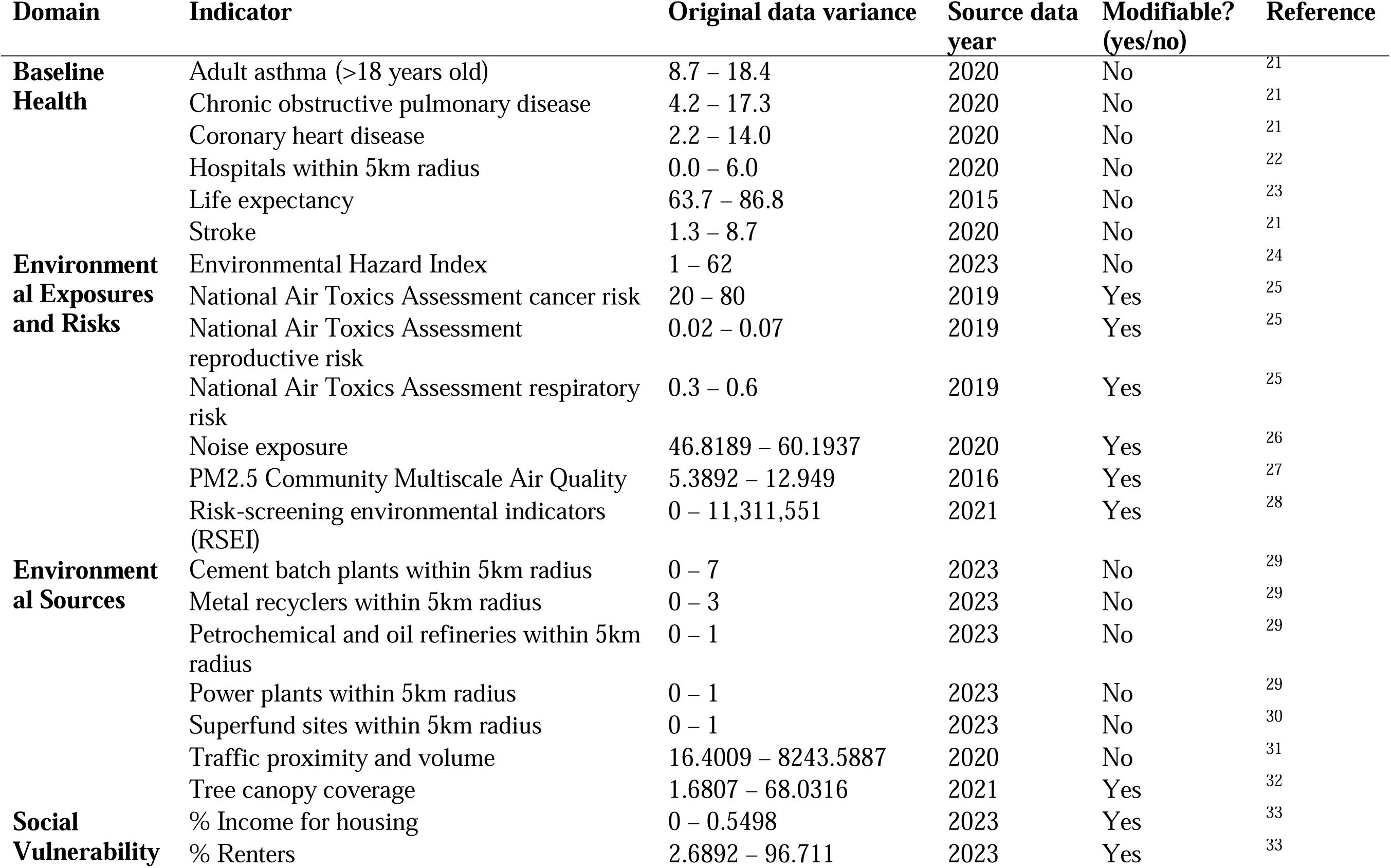
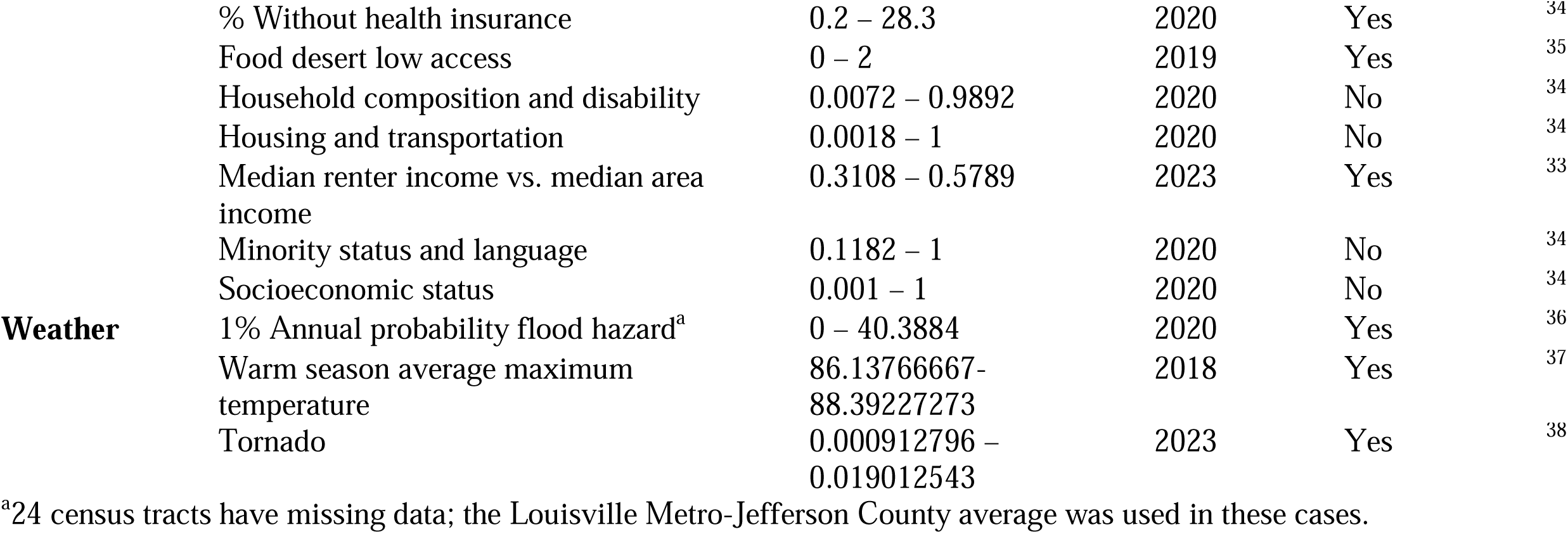
Domains and vulnerability Indicators in the Environmental Vulnerability Index model for Louisville Metro-Jefferson County, Kentucky (USA), data variance, source dataset publication year, and assessment of if modification is possible by public health intervention or policy.

The baseline health domain contains six indicators pertaining to disease, longevity, and healthcare access to show where residents themselves are most vulnerable to adverse environmental impacts. The environmental exposures and risks domain encompasses nine indicators related to ambient pollution and general environmental hazards. The environmental sources domain features seven indicators, six reflect point sources of environmental exposures and the seventh is tree canopy. The social vulnerability domain includes nine indicators related to the resilience capacity of communities to recover from natural disasters and other crises. The extreme weather domain includes three indicators: flood, heat exposure, and tornados.

### Data Analysis

The Toxicological Prioritization Index (ToxPi) version 1.2.1 (Durham, North Carolina) was used to produce EVI scores by census tract (n=190).^3,4,39^ Indicators were converted to percentiles to put them on the same scale before being combined within domains. Equal weights were applied to each indicator within a domain, and each domain was weighted equally as one-fifth. Scores are relative rankings; a composite score of 0 indicates that an area has no vulnerabilities while higher scores indicate more vulnerability. The Louisville International Airport comprises an entire census tract and was excluded. Maps, geocoding, and Local Moran’s I analysis were performed using ArcGIS Pro version 2.9.5 (Redlands, CA).

### Environmental Vulnerability Intervention Simulation

To evaluate potential public health interventions for reducing environmental vulnerabilities, modifiable indicators were identified, and three intervention scenarios were developed (Figure 2). Indicators were determined to be modifiable if they were changeable by policy or other intervention activity within five years, for instance – tree planting efforts can increase the percentage of tree canopy and air toxics can be reduced through policy or power plant retirement, retrofit, and conversion to natural gas.^7,40^ To define modifiable factors, particular emphasis was placed on their potential for theoretically feasible implementation in real-world settings,^41^ within the context of census tract-level influence. Non-modifiable indicators included factors such as interstate highways, industrial corridors, rivers, and other natural landscape features.

**Figure 2.**
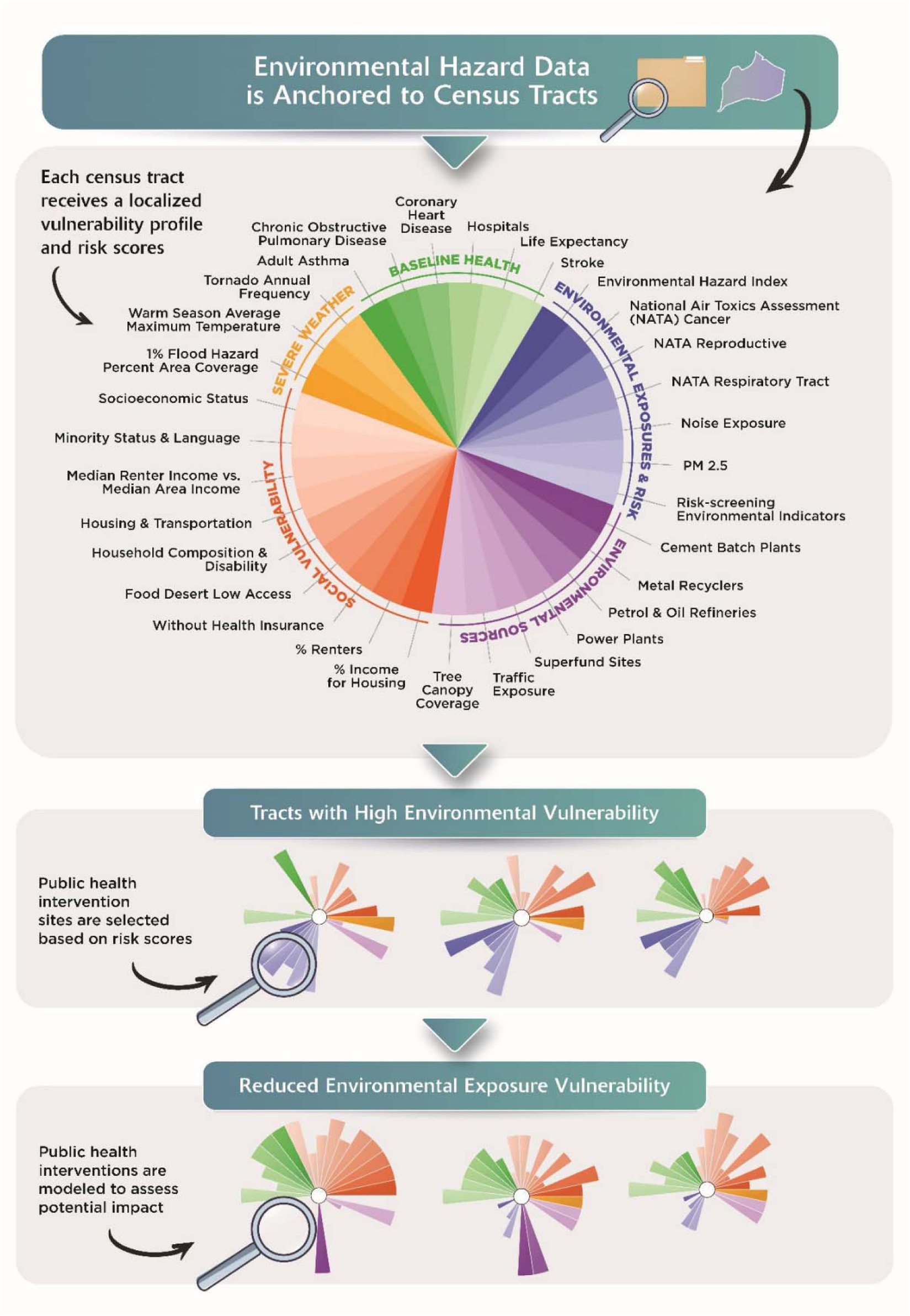
Framework for Environmental Vulnerability Index supporting simulation of targeted public health interventions at the census tracts level. Using the Toxicological Prioritization Index composite scores, the five most vulnerable census tracts were identified and modifiable indicators were selected under three intervention scenarios as theoretical action for solutions.

The five most vulnerable census tracts, those with the highest ToxPi composite scores, were selected as intervention sites with three illustrative case studies for each (A, B, C). To model interventions, we substituted selected indicator score(s) with the lowest (least vulnerable) score observed in Louisville Metro-Jefferson County. This change represents a theoretical intervention aimed at improving the tract’s overall vulnerability. Intervention A modeled the potential impacts improving singular indicators such as tree canopy, noise pollution exposure, or respiratory risk from air toxics. Intervention B modeled the potential impacts of improving an entire domain such as environmental exposures and risks, environmental sources, or weather; for example, an intervention that included removing a group of point sources of pollution, decreasing traffic and increasing tree canopy coverage through joint economic and policy investment. Intervention C modeled the potential impacts of improving a thematic cluster of indicators, such as air pollution, environmental infrastructure, or point sources of pollution (Table S4).

### Ethics

Data used in the analysis are available in online public records, sources are provided in Table S2.

## Results

### Composite ToxPi Scores

Composite ToxPi scores were not uniformly distributed across Louisville Metro-Jefferson County (Figure 3); they ranged from 0.19 to 0.71. Geographically, high vulnerability scores were observed along the western and central areas of the county with lower vulnerability scores in the central urban core and eastern regions. The most at-risk census tract (21111005900) is in the downtown urban core. The least at-risk census tract (21111013100) is a mainly residential area in the urban core that abuts a park and a local airfield. However, there are pockets of vulnerability and census tracts that span geographic locations. The five least vulnerable census tracts are spread across an area east of Interstate 65. In contrast, the five most vulnerable census tracts are spread across the central and western areas of the county. This pattern matches the distribution of CEJST-designation of disadvantaged.^20^ Of the 72 disadvantaged census tracts in Jefferson County 83% (70) are in the western and south-central regions. Of the 21 most vulnerable census tracts, 80% (17) are considered disadvantaged. All five of the most vulnerable census tracts from our model are also considered CEJST disadvantaged.

**Figure 3.**
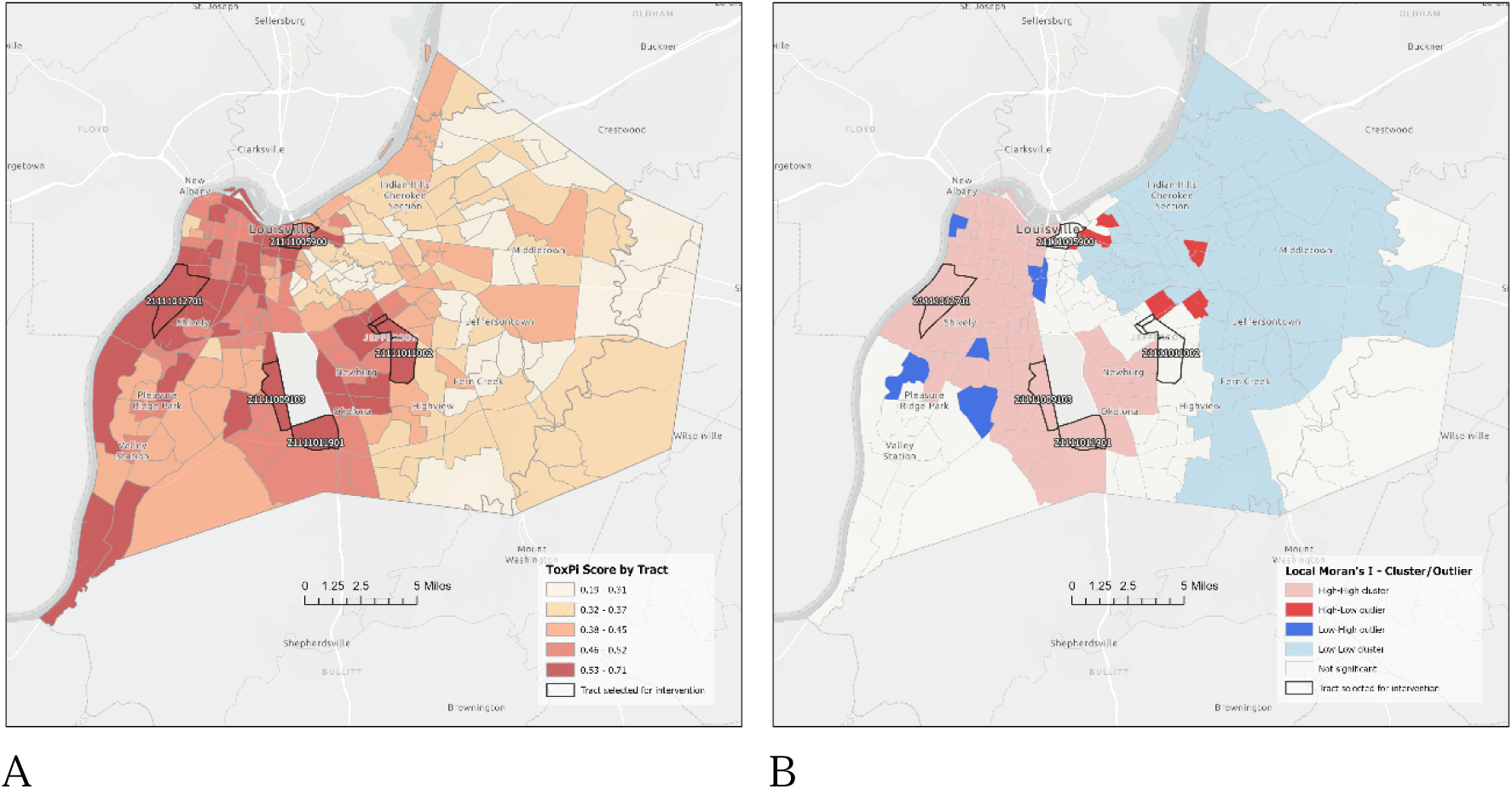
Toxicological Prioritization Composite Score Index by census tract, Louisville Metro-Jefferson County, Kentucky (USA). Panel A: Composite scores by census tract whereby a higher score corresponds to a more vulnerable census tract. Census tracts were sorted into quartiles by overall score, the quartiles were scored as: 1^st^ quartile as lowest vulnerable census tracts <0.32 (n=48); 2^nd^ quartile 0.33 to 0.41 (n=48); 3^rd^ quartile 0.42 to 0.51 (n=47); and 4^th^ quartile as most vulnerable census tracts >0.52 (n=47). Panel B: Modeled clusters of composite scores by census tract. The Local Moran’s I analysis indicates statistically significant clusters of high or low values, as well as outliers where high values are adjacent to low values, and vice-versa.

Local Moran’s I analysis was used to identify statistically significant clusters and outliers. Clusters are areas where tracts with high or low ToxPi scores are adjacent to other tracts with similarly high or low scores. Outliers are areas where tracts with high values are adjacent to low values and vice-versa (Figure 3). Results show significant clustering of higher ToxiPi scores in the west, with low ToxiPi scores cluster in the eastern county areas. There are five outlier tracts with relatively higher ToxPi scores than their neighboring tracts in the eastern county areas.

### Domain-specific Scores

Some census tracts with low composite ToxPi scores have high domain-specific scores (Figure 4). The five most vulnerable census tracts especially score low in the domains of environmental exposures and risks and environmental exposures. The least vulnerable census tract (21111013100) has the best score across the county for both for social vulnerability and severe weather domains.

**Figure 4.**
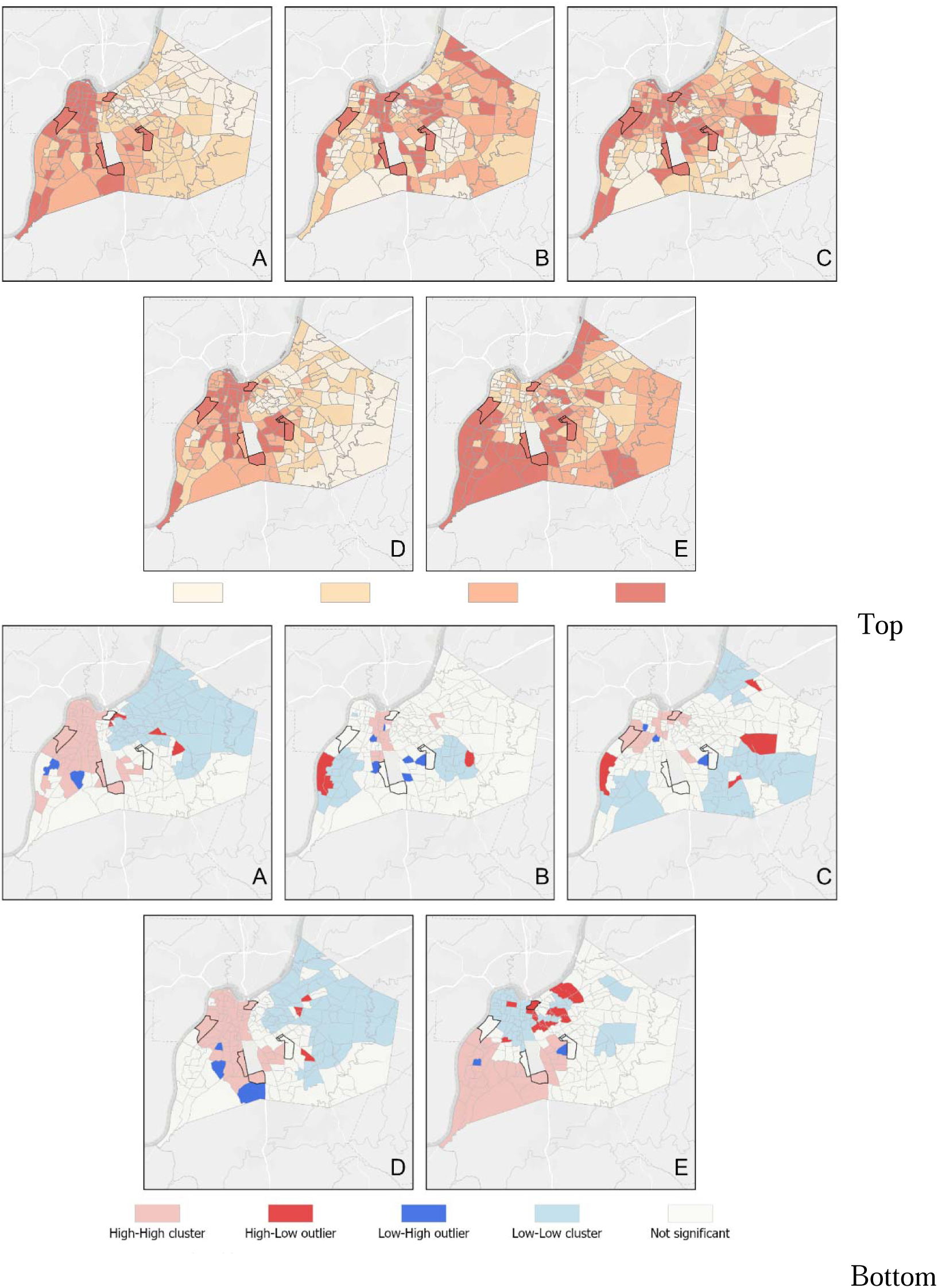
Toxicological Prioritization Domain Score Index by census tract, Louisville Metro-Jefferson County, Kentucky (USA). Top: Domain scores by census tract whereby a higher score corresponds to a more vulnerable census tract. Panels present the domain scores for: A.) Baseline Health; B.) Environmental Exposures and Risks; C.) Environmental Sources; D.) Social Vulnerability; and E.) Extreme Weather. Bottom: Modeled clusters of domain scores by census tract. The Local Moran’s I analysis indicates statistically significant clusters of high or low values, as well as outliers where high values are adjacent to low values, and vice-versa. Panels present the domain scores for: A.) Baseline Health; B.) Environmental Exposures and Risks; C.) Environmental Sources; D.) Social Vulnerability; and E.) Extreme Weather.

### Heterogeneity

Heterogeneity analysis (Figure S1) was performed for each for each of the 32 indicators to ensure only factors that offer significant variance were included in the index. Indicators that have limited variation thus may have minimal influence on the model’s predictive accuracy. Heat exposure and noise are two examples with a good, but narrow, data range across census tracts. As well, for NATA cancer, most (186/190) census tracts have the same raw value, but there is some variation. And, again for renter/owner income most (185/190) census tracts have the same raw value. If the framework indicated an indicator raw score that had no-variation it would have been appropriate to remove; no changes were made following heterogeneity analysis.

### Modelling the Impact of Interventions on Vulnerability Scores

Across the Intervention A case studies, modifications to a single indicator led to enhancements in the ToxPi composite score ranging from 1 to 4% (Table 2). Improving single indicators does not universally reduce vulnerability, even in the most vulnerable tracts. Improving the best possible indicator score reflected in the county for tree canopy alone resulted in an average 3% improvement to the composite ToxPi score, while improving noise pollution or NATA respiratory risk were an average 2% improvement. Currently tree canopy coverage is as low as 5% and adjusting tree canopy in these five census tracts uniformly to 10% only improved composite ToxPi scores marginally, an average of 1% improvement. Increasing tree canopy to the countywide goal of 45%^42^ improved average composite ToxPi scores by 3%. One of the most vulnerable census tracts (21111011901) already had a 31% tree canopy coverage, which is good for an urban area and better than the average tree canopy coverage across the county.

**Table 2.**
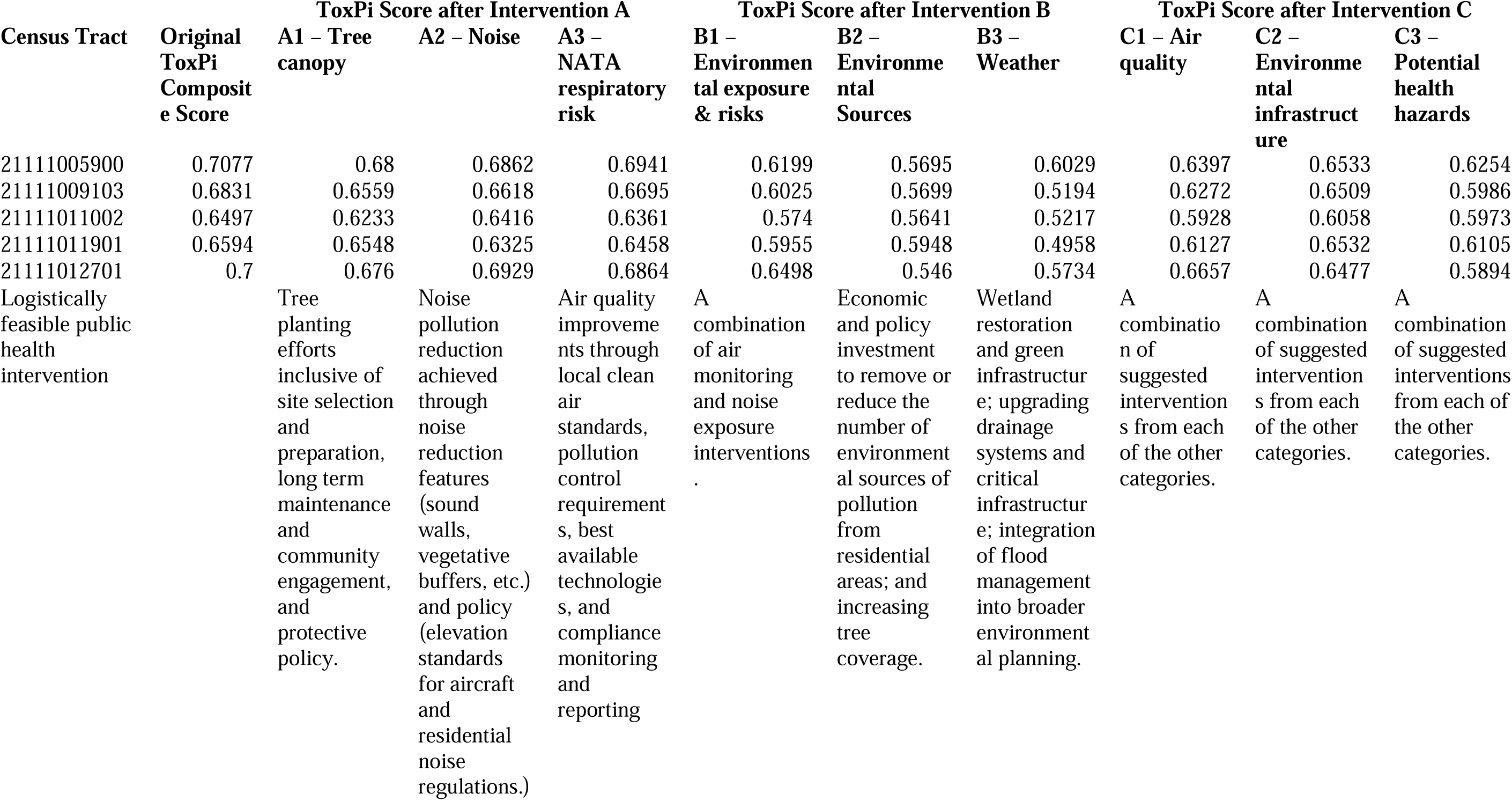
Toxicological Prioritization Index (ToxPi) composite scores for the lowest five ranking census tracts and modeled case study result following intervention to the best possible indicator score reflected in the county. Higher composite scores correspond to higher vulnerability. Intervention A modeled the potential impacts of improving one indicator while keeping all other indicators static. Intervention B modeled the potential impacts of improving entire domains. Intervention C modeled the potential impacts of improving clusters of indicators with related interventions. Each model intervention was conducted three times as an illustrative case studt.

| Census Tract | Original ToxPi Composite Score | ToxPi Score after Intervention A |  |  | ToxPi Score after Intervention B |  |  | ToxPi Score after Intervention C |  |  |
| --- | --- | --- | --- | --- | --- | --- | --- | --- | --- | --- |
|  |  | A1 – Tree canopy | A2 – Noise | A3 – NATA respiratory risk | B1 – Environmental exposure & risks | B2 – Environmental Sources | B3 – Weather | C1 – Air quality | C2 – Environmental infrastructure | C3 – Potential health hazards |
| 21111005900 | 0.7077 | 0.68 | 0.6862 | 0.6941 | 0.6199 | 0.5695 | 0.6029 | 0.6397 | 0.6533 | 0.6254 |
| 21111009103 | 0.6831 | 0.6559 | 0.6618 | 0.6695 | 0.6025 | 0.5699 | 0.5194 | 0.6272 | 0.6509 | 0.5986 |
| 21111011002 | 0.6497 | 0.6233 | 0.6416 | 0.6361 | 0.574 | 0.5641 | 0.5217 | 0.5928 | 0.6058 | 0.5973 |
| 21111011901 | 0.6594 | 0.6548 | 0.6325 | 0.6458 | 0.5955 | 0.5948 | 0.4958 | 0.6127 | 0.6532 | 0.6105 |
| 21111012701 | 0.7 | 0.676 | 0.6929 | 0.6864 | 0.6498 | 0.546 | 0.5734 | 0.6657 | 0.6477 | 0.5894 |
| Logistically feasible public health intervention |  | Tree planting efforts inclusive of site selection and preparation, long term maintenance and community engagement, and protective policy. | Noise pollution reduction achieved through noise reduction features (sound walls, vegetative buffers, etc.) and policy (elevation standards for aircraft and residential noise regulations.) | Air quality improvements through local clean air standards, pollution control requirements, best available technologies, and compliance monitoring and reporting | A combination of air monitoring and noise exposure interventions. | Economic and policy investment to remove or reduce the number of environmental sources of pollution from residential areas; and increasing tree coverage. | Wetland restoration and green infrastructure; upgrading drainage systems and critical infrastructure; integration of flood management into broader environmental planning. | A combination of suggested interventions from each of the other categories. | A combination of suggested interventions from each of the other categories. | A combination of suggested interventions from each of the other categories. |

For Intervention B, entire domains with several indicators were adjusted: environmental exposures and risks, environmental sources, or weather. Across the case studies, domain modifications led to enhancements in the ToxPi composite scores ranging from 7% to 25%. When indicator scores within a domain were adjusted to the best scores observed in the county, the census tract’s composite ToxPi scores were reduced by an average of 11% for environmental exposures and risks, 16% for environmental sources, and 20% for extreme weather.

For Intervention C, thematically related intervention clusters of indicators were adjusted: air pollution, environmental infrastructure, or potential health hazards. Across the case studies, modifications to these clusters led to enhancements in the ToxPi composite scores ranging from 1 to 16%. When indicator scores for each cluster were reduced to the best scores observed in the county, the census tract’s composite ToxPi scores were reduced on average for air quality at 8%, for environmental infrastructure at 6%, and for potential health hazards at 11%.

## Discussion

In this study, we used a modified version of community based EVI framework^1^ to assess vulnerabilities in a new geographic region. We extended the tool to model interventions and to assess potential impact on hyperlocal risks. Our approach provides a data-driven guide for Louisville Metro-Jefferson County as examples of census tract scale solutions as opposed to more general public health programming. The index is customized to the particular concerns of our area, including the addition of indicators such as noise pollution exposure, heatwave exposure, and tornados. While Messer et al.^43^ developed neighborhood socioeconomic context for 19 cities, it excluded flood, growing urban heat island effect, and tornados which are locally important vulnerabilities in the Louisville Metro-Jefferson County area. The presence of a commercial air-hub led to the addition of the noise pollution indicator. These risks are real for many cities and the expanded evaluation demonstrates the benefits of adaptation to local conditions, risks, and vulnerabilities. The Louisville Metro-Jefferson County has unique vulnerabilities when compared with most other counties across of the nation, although the area does resemble several towns in the United States Midwest. For example, all 190 census tracts in Louisville Metro-Jefferson County have higher average PM 2.5 concentrations than the nationwide average of 3.79 µg/m3. Identification and incorporation of such local vulnerabilities may be key to developing local and well-targeted interventions.

Previous work suggests that single-indicator interventions to improve health are theoretically feasible. Wang et al.^40^ reported on the effectiveness of an urban green and blue space intervention in improving wellbeing. Brown et al.^44^ and Hammer et al.^45^ discovered that direct regulation on lowering noise at its source, expanding and increasing access to noise maps, and altering the built environment can be effective. Further, Casey et al.^7^ documented improved health outcomes resulting from the removal of an environmental point source of pollution when the retirement of a coal-fired power plant led to improved asthma outcomes.

Our index integrates several related indicators into a single domain and it incorporates social, clinical, and environmental data to provide a more comprehensive evaluation of area vulnerability. In previous work, other environmental health vulnerability studies conducted at census tract level have focused on only clinical data or only a few public data sources,^40,46^ despite the probability that a wide range of structural environmental variables can impact health. For instance, social vulnerability combines data on socioeconomic status, housing and transportation, and health insurance coverage. When assessing access to healthcare, this combined view of indicators may be more explanatory of lack of healthcare access than a simple hospital proximity indicator.

Our findings suggest that environmental vulnerability could be feasibly estimated at census tract scale, rather than county level, consistent with the Glassman et al.^5^ report. In addition, our work demonstrates that county-level data could be too broad to support intervention evaluation. Census tract level data offers balanced spatial granularity useful to discern hyperlocal variations within urban areas whereby environmental risks can be viewed within the context of the entire spectrum of risks.

The index enabled the selection of the most at-risk census tracts for modeling targeted public health interventions to reduce cumulative environmental vulnerability within specific communities. Our work also suggests that while some census tracts may require plans tailored to those specific areas, it may be possible to develop interventions effective in addressing common problems that span boundaries across multiple tracts. Moreover, our work across the three case studies also suggests that improving any single indicator may not be highly beneficial in addressing the overall area vulnerability and that it may be necessary to address multiple indicators collectively. Interesting, a high composite vulnerability scores can mask low indicator-level vulnerabilities (e.g., good tree coverage). Overall, our index and our method for computing aggregate vulnerabilities maybe helpful in identifying focal points for resource allocation. However, additional work will be required to assess the efficacy of any intervention. For instance, Louisville’s 14 hospitals are clustered in one geographic area thus limiting access, as measured by geographical proximity, for many census tracts. Although this proximity indicator reflects an equal impact geographically, it may result in a stronger impact on individuals at the low end of the socioeconomic spectrum compared to those at the highest; affluent individuals have significant resources, personal transportation, and insurance to access healthcare, regardless of distance to the nearest hospital. Thus, a healthcare access intervention may be warranted for low-income census tracts to offset this inequity. Moreover, in our area of interest seven census tracts are within a 5-kilometer radius of Superfund sites, and all of these are west of a structural environment variable, Interstate 65, mirroring larger vulnerability patterns in the county. This suggests purposively modifying some indicators may have a disproportionately positive impact on the county’s most vulnerable census tracts. Therefore, as indicated by these two examples, several indicators are highly correlated and therefore changes in one could have far reaching effect on the other or may benefit only related aspects of those vulnerabilities.

Threshold values across both the composite score and indicators can be used to manage expectations related to interventions. More affluent census tracts in Jefferson County have ToxPi composite scores of less than 0.3. These areas are often characterized by high life expectancy, low social vulnerability, and limited pollution exposure. As well per a single indicator, while 40-45% is a goal set by Louisville’s Urban Tree Canopy Assessment^42^ the lowest tree canopy score in our study area is 2%, and any short-term aim to increase the canopy indicator to 40% may be a highly unrealistic goal. When we modeled the 45% canopy coverage, it only improved the ToxPi composite score to be 3% better. These results put in perspective the expected indicator gains that could be accrued by specific interventions and should help in prioritizing interventions. Moreover, some indicator scores may not change without a modification in direct policy and/or financial intervention, such as the presence of industrial facilities. Finally, when selecting areas for an intervention, it may be important to consider ethical questions around withholding environmental health interventions known to be effective.^47^

The main purport of the framework developed by our work may be to guide actions to mitigate vulnerability and risk. For example, local governance could use this framework to create action plans for increasing neighborhood resilience and preparedness following major events. Local public health agencies could use also digital platforms and targeted advertising campaigns to raise awareness and promote health actions in specific places.^48^ Qualitative researchers could use the vulnerability index to gather community input to validate proposed solutions. Finally, grassroots community organizations could use an assessment of environmental vulnerability to identify problems, construct potential solutions, and advocate for policy change. Establishing a data-driven index to inform interventions tailored to specific needs, rather than generic approaches would also ensure accountability by verifying that the interventions are directed towards areas where they are most needed and address the area’s most significant vulnerabilities.

Overall, we believe our study advances the integrated vulnerability index approach for selecting census tracts for health interventions in several key ways:

1. **Consideration of multiple domains and indicators**. Consideration of health parameters beyond traditional clinical and environmental data can inform more effective, data-driven, public health communications, and interventions.
2. **Data anchored to census tract boundaries**. Focus on census tract level vulnerability rather than countywide scale.
3. **Prioritization of intervention and investment by data-driven vulnerability**. Our approach shows which census tracts are in most need of vulnerability mitigation and intervention. In some cases, the same intervention may benefit multiple census tracts even if they are not geographically adjacent.
4. **Shift focus away from the single indicator interventions**. Modeled results show improving a group of related indicators is more impactful than adjusting a single indicator.

Despite its many strengths, the study has some limitations. We relied solely on publicly available national, state, and local government data sources, potentially overlooking data requiring Data Transfer Agreements or open records requests. Application may be limited for rural census tracts due to a lack of local data available. We restricted our analysis to Louisville Metro-Jefferson County, neglecting the bordering counties of the metro area. All domains are equally weighted as we could not justify indicator-specific weighting. Qualitative research and community input can be used in tools to validate vulnerability frameworks; however, integration of qualitative and quantitative data sources can pose additional challenges.^49^ Our modeled interventions have limitations; determining efficacy of intervention requires empirical testing to establish the degree to which environmental vulnerabilities are modifiable. One major limitation of our approach is that scores are percentile-based which does not distinguish between indicators with low and high variance. However, we addressed this limitation by selecting indicators for interventions that could feasibly produce a material change, such as percent tree canopy, achievable in real-world conditions.

## Conclusion

In this study we used census tract boundaries and integrated vulnerability data to develop a comprehensive vulnerability index which could be used to assess the potential impact of targeted public health interventions. While the original framework was designed to assign relative ranks based on environmental vulnerability, the aggregation of vulnerability and intervention allows the index to develop into a more comprehensive tool for public health, community planning, and environmental justice advocates. Chief among our insights is that places are not defined by their environmental vulnerabilities and that such vulnerabilities are not invariant features of living communities. Rather, knowledge of these risks can spur action for towards local and bespoke solutions. Targeted census tract interventions may be more effective than broad-scale campaigns at state or national levels and may lead to health-supportive environmental conditions with greater geographic precision.

## Data Availability

All source data are publicly available, links are provided in Table S2.

## Contributors

Conceptualization: TS, WC, AB; Methodology: TS, AB; Formal analysis: LBA, CB, DJB, RHH; Writing-original draft preparation: LBA, RHH; Writing-review and editing: TS, LBA, CB, DJB, WC, RHH, AB; Supervision: TS, AB; Project administration: LBA. All the authors have read and agreed to the published version of the manuscript.

## Data sharing

All source data are publicly available, links are provided in Table S2.

## Acknowledgments

The authors thank Ray Yeager, Rebecca Turney, and Angelina Rangel for their mapping expertise, Dhiraj Kanneganti for ToxPi skills developed during this project, and Laney Taylor for gathering and organizing the initial data for this research.

## Supplementary Information

**Table S1.**
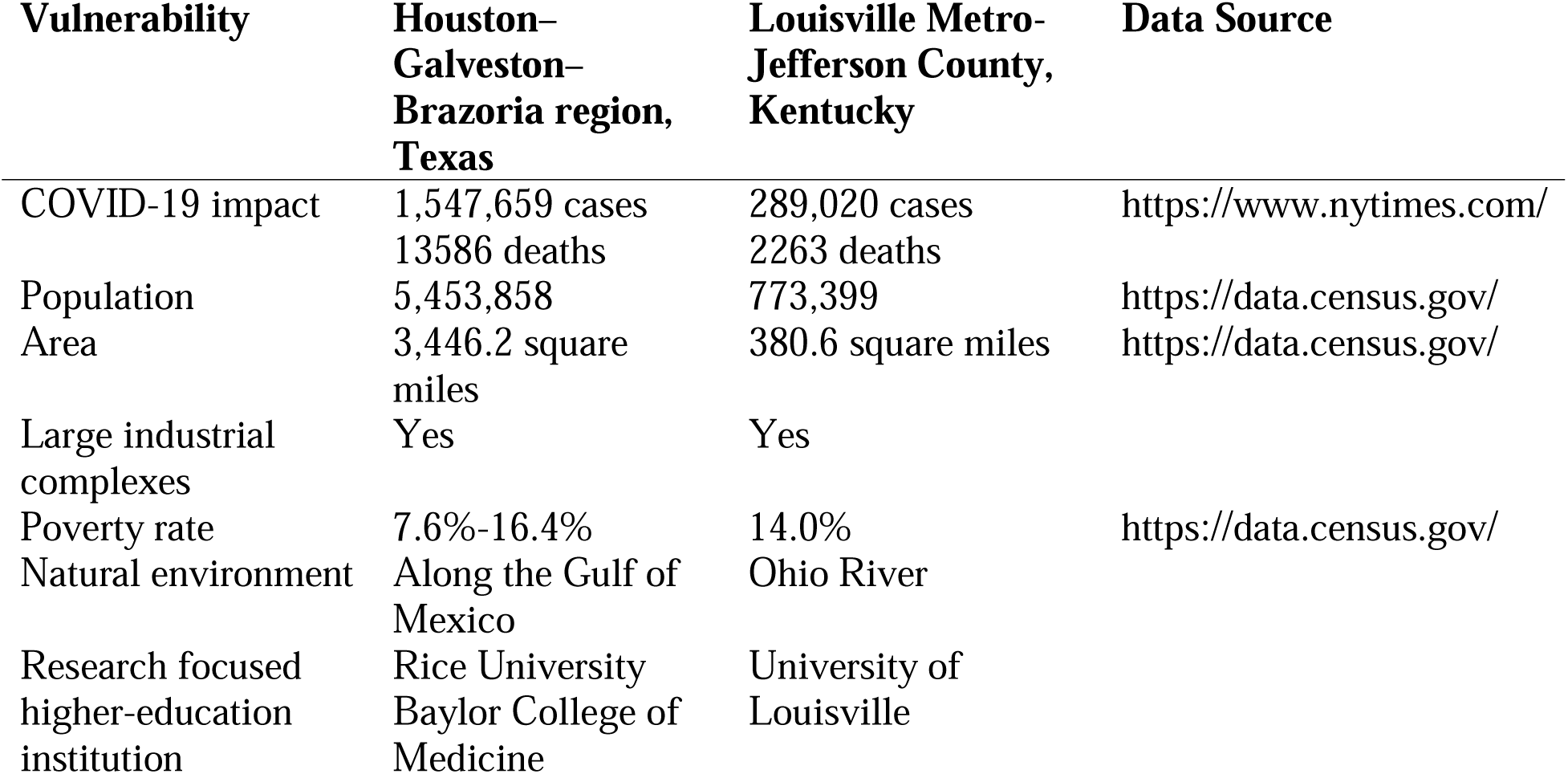
Vulnerabilities of Houston–Galveston–Brazoria region, Texas, and Louisville Metro-Jefferson County, Kentucky.

| **Vulnerability** | **Houston–Galveston–Brazoria region, Texas** | **Louisville Metro-Jefferson County, Kentucky** | **Data Source** |
| --- | --- | --- | --- |
| COVID-19 impact | 1,547,659 cases<br>13586 deaths | 289,020 cases<br>2263 deaths | https://www.nytimes.com/ |
| Population | 5,453,858 | 773,399 | https://data.census.gov/ |
| Area | 3,446.2 square miles | 380.6 square miles | https://data.census.gov/ |
| Large industrial complexes | Yes | Yes |  |
| Poverty rate | 7.6%-16.4% | 14.0% | https://data.census.gov/ |
| Natural environment | Along the Gulf of Mexico | Ohio River |  |
| Research focused higher-education institution | Rice University<br>Baylor College of Medicine | University of Louisville |  |

**Table S2.**
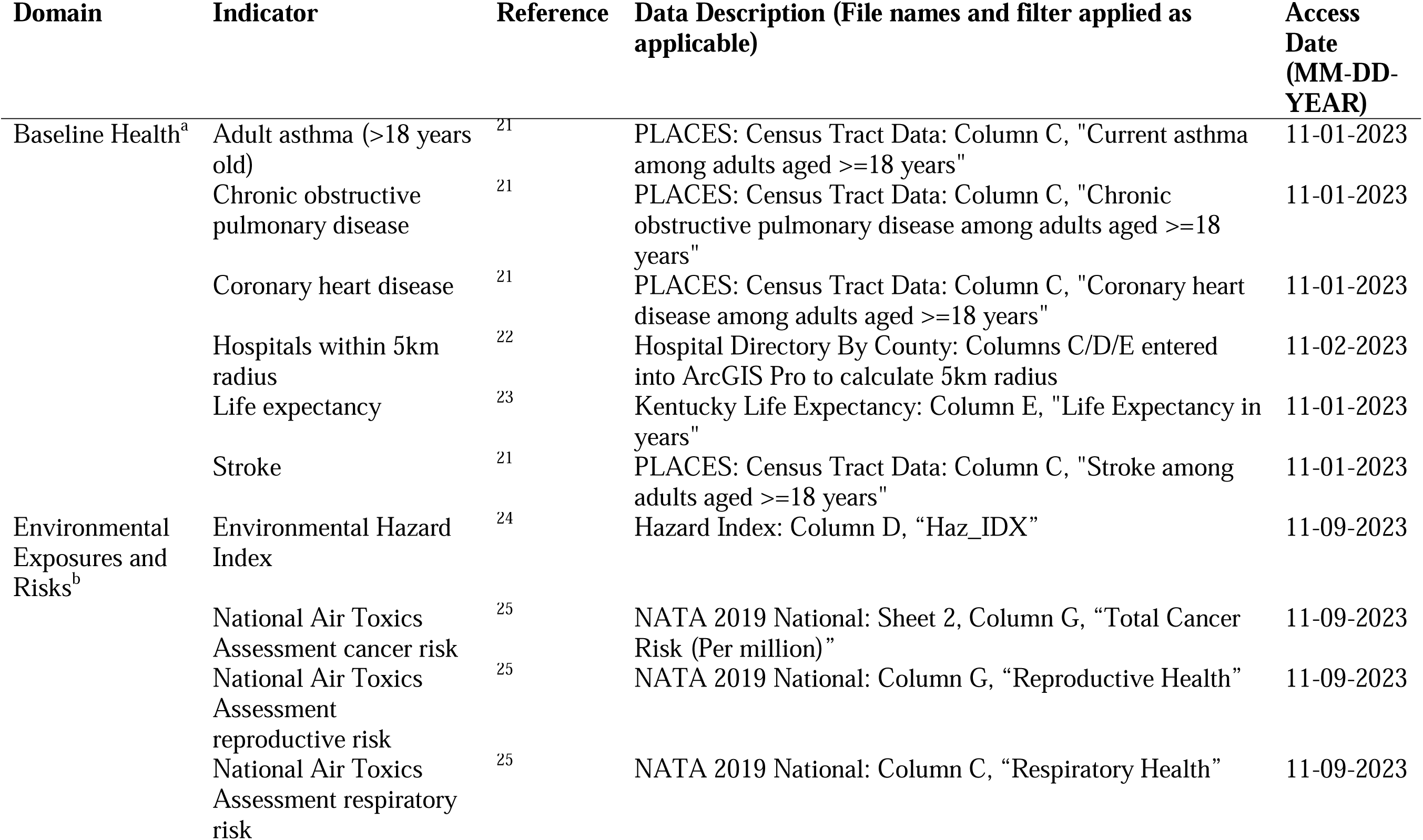
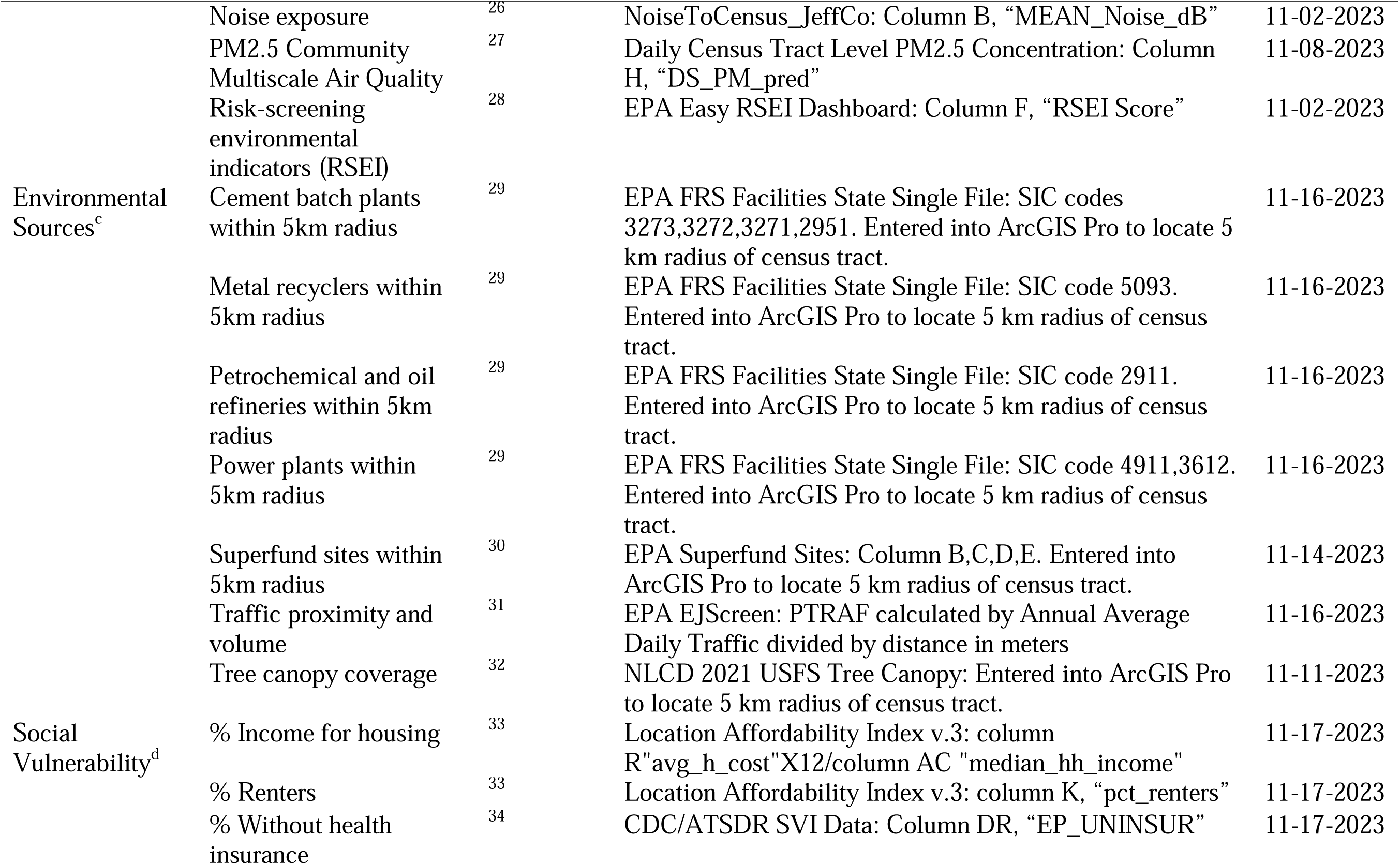
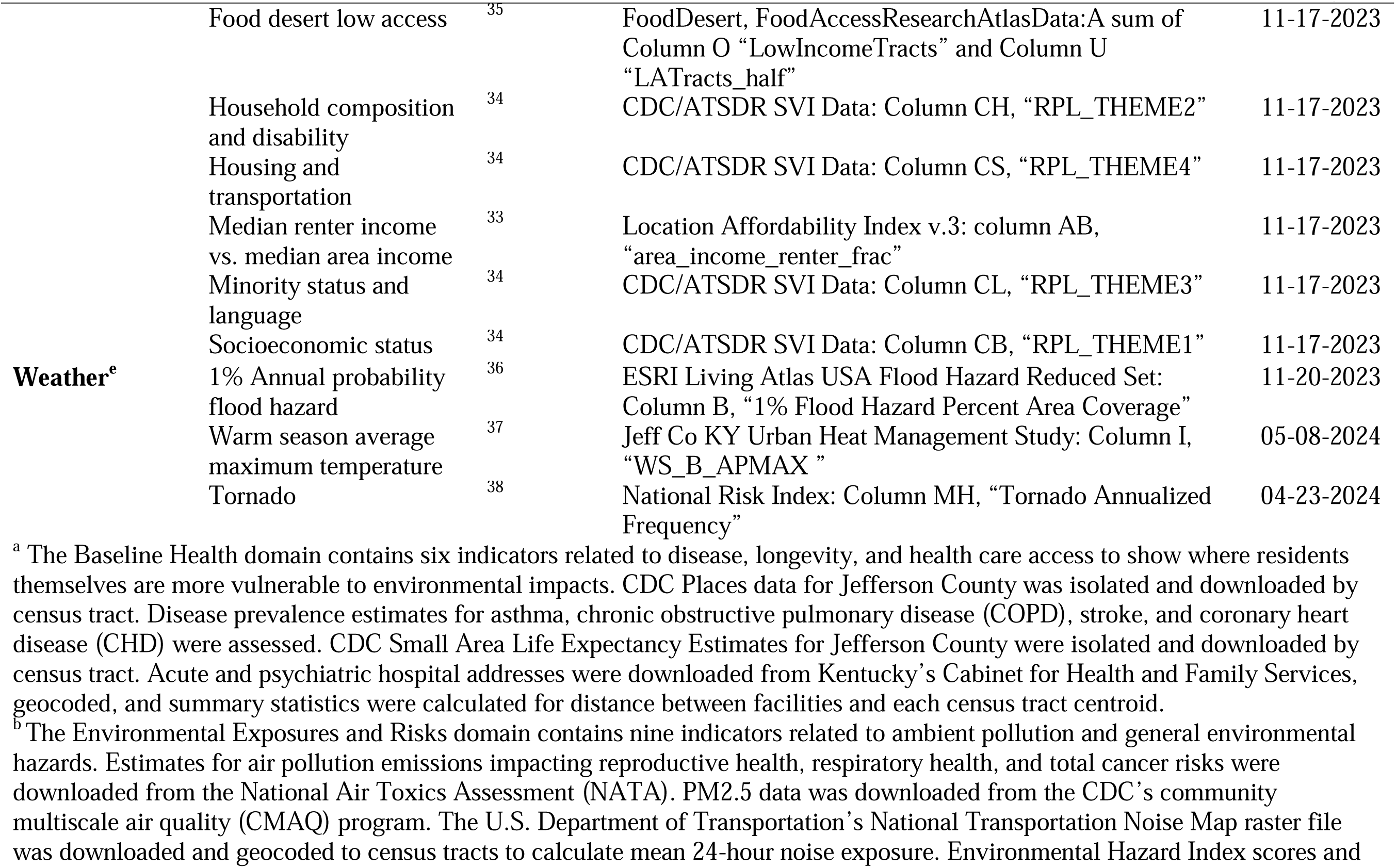
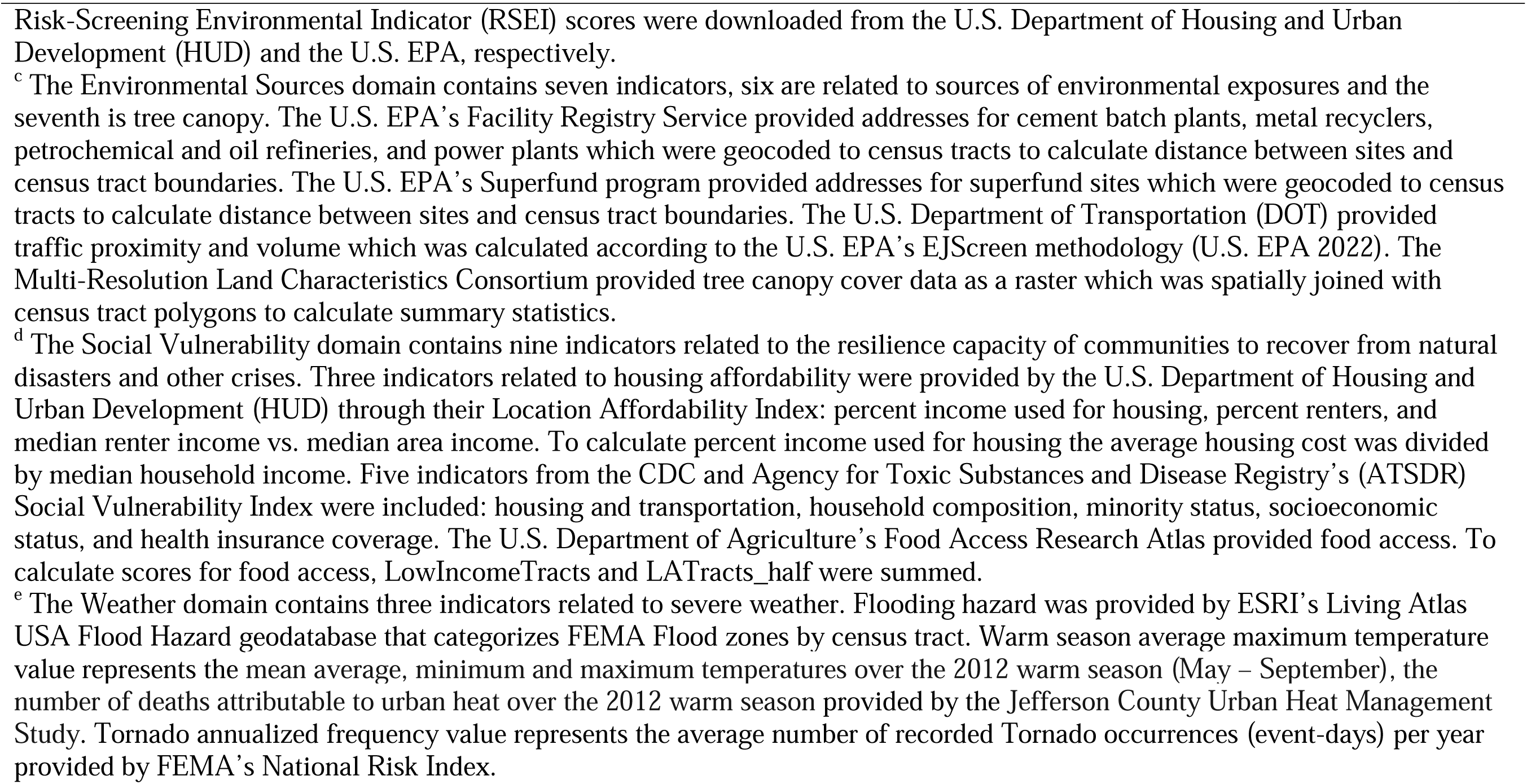
Data sources of indicators incorporated into Environmental Vulnerability Index for Louisville Metro-Jefferson County, Kentucky (USA).

**Table S3.**
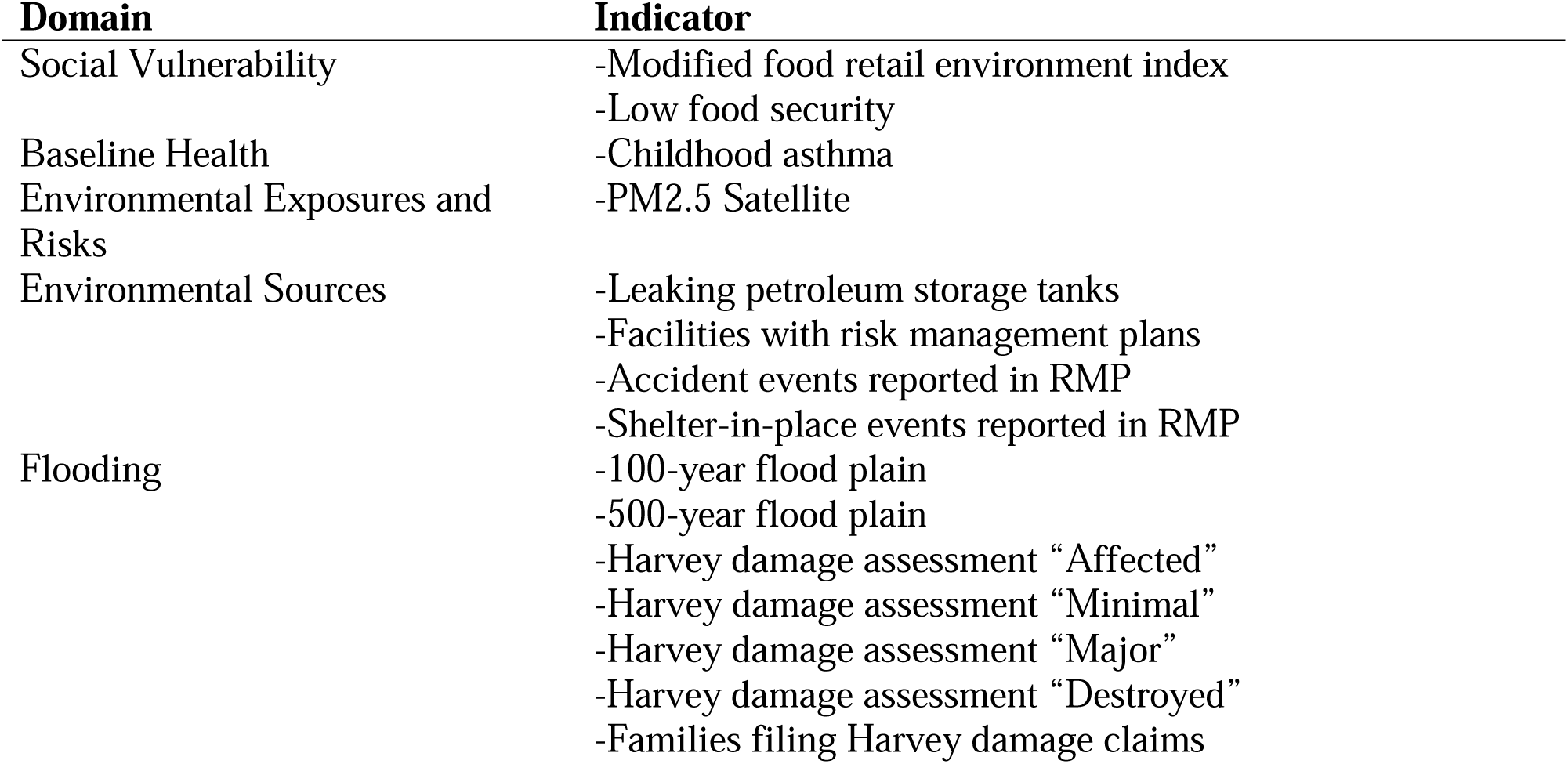
Indicators included in the Houston–Galveston–Brazoria (HGB) EnviroScreen’s Environmental Vulnerability Index (EVI) (Bhandari et al., 2020) which were removed for the Louisville Metro-Jefferson County index.

| **Domain** | **Indicator** |
| --- | --- |
| Social Vulnerability | -Modified food retail environment index<br>-Low food security |
| Baseline Health | -Childhood asthma |
| Environmental Exposures and Risks | -PM2.5 Satellite |
| Environmental Sources | -Leaking petroleum storage tanks<br>-Facilities with risk management plans<br>-Accident events reported in RMP<br>-Shelter-in-place events reported in RMP |
| Flooding | -100-year flood plain<br>-500-year flood plain<br>-Harvey damage assessment “Affected”<br>-Harvey damage assessment “Minimal”<br>-Harvey damage assessment “Major”<br>-Harvey damage assessment “Destroyed”<br>-Families filing Harvey damage claims |

**Table S4.**
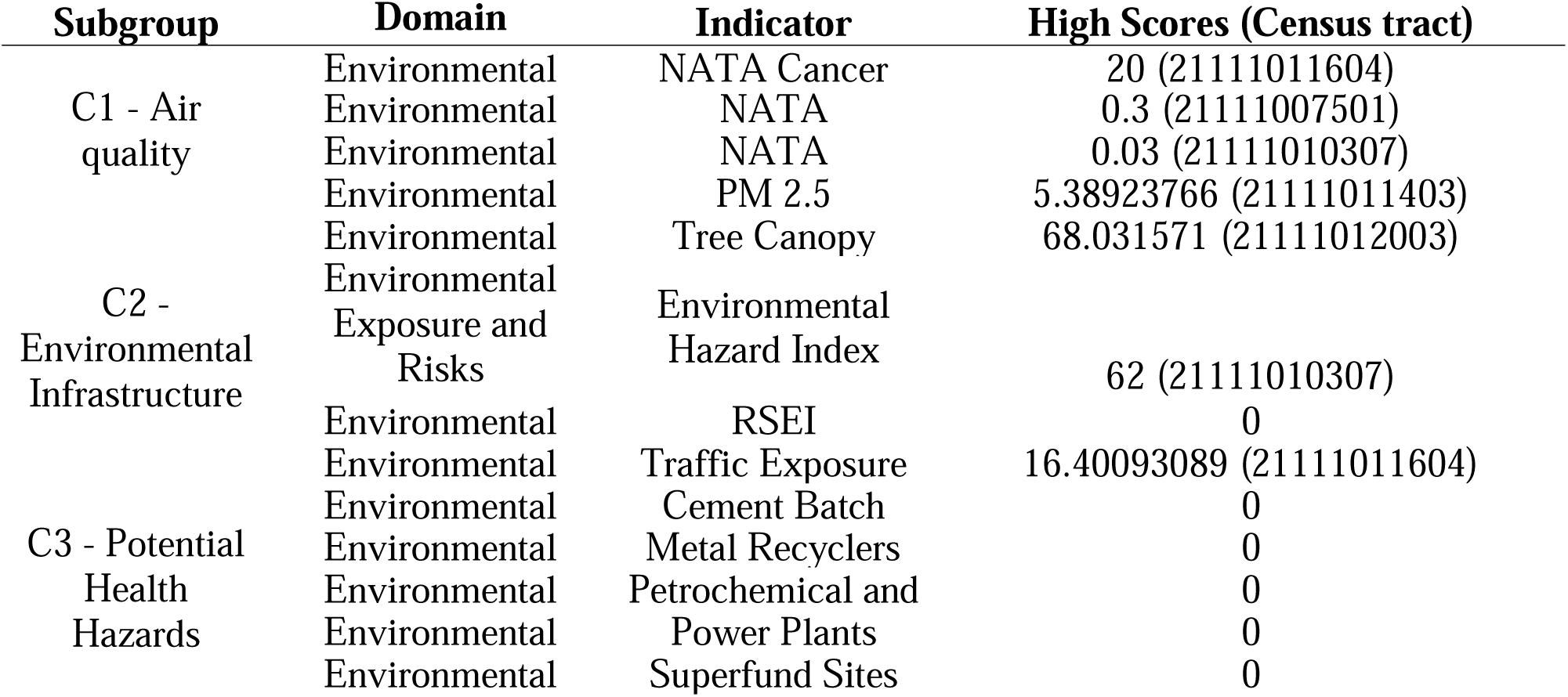
Indicators subgrouped based upon related impacts across domains for Intervention C case studies.

**Figure S1.**
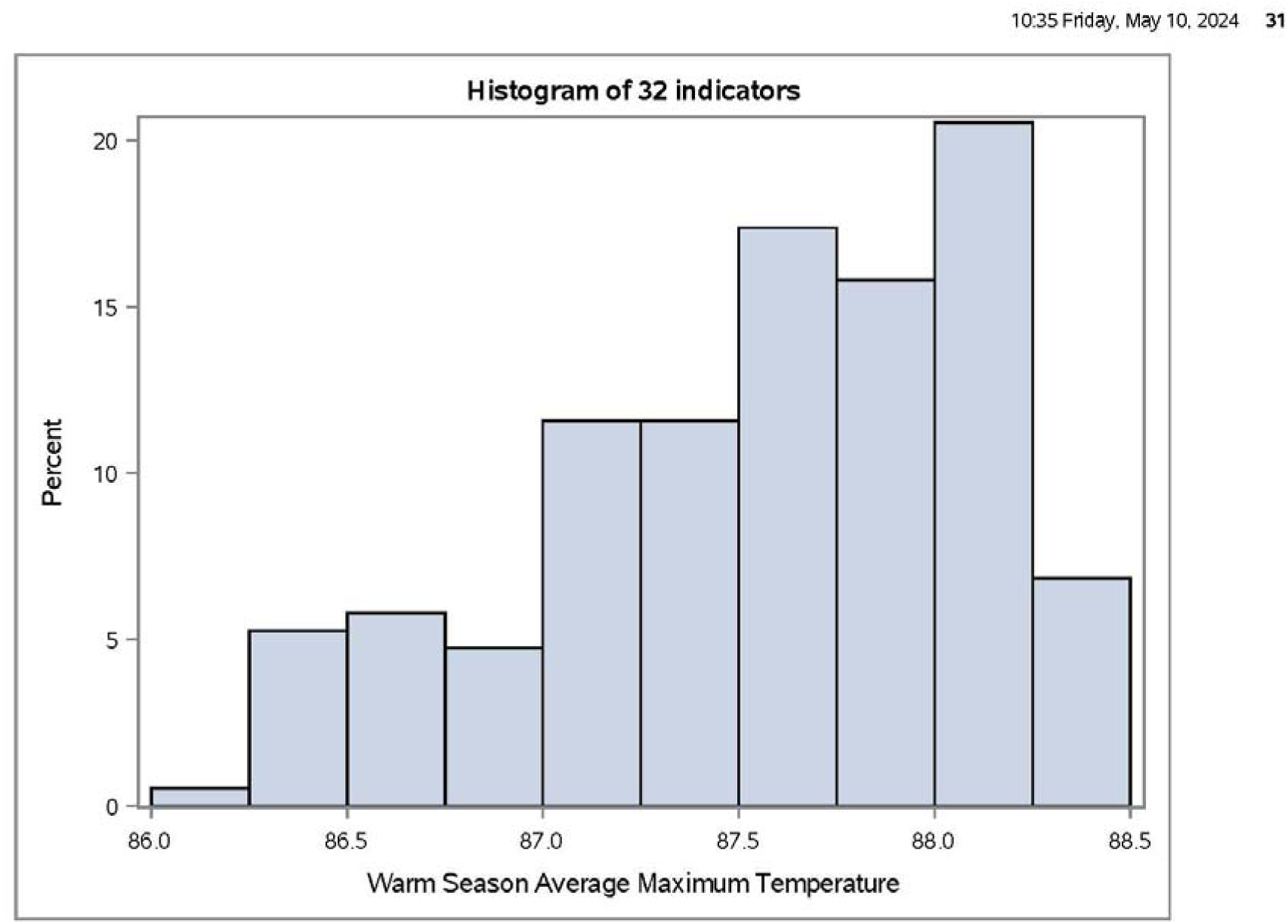
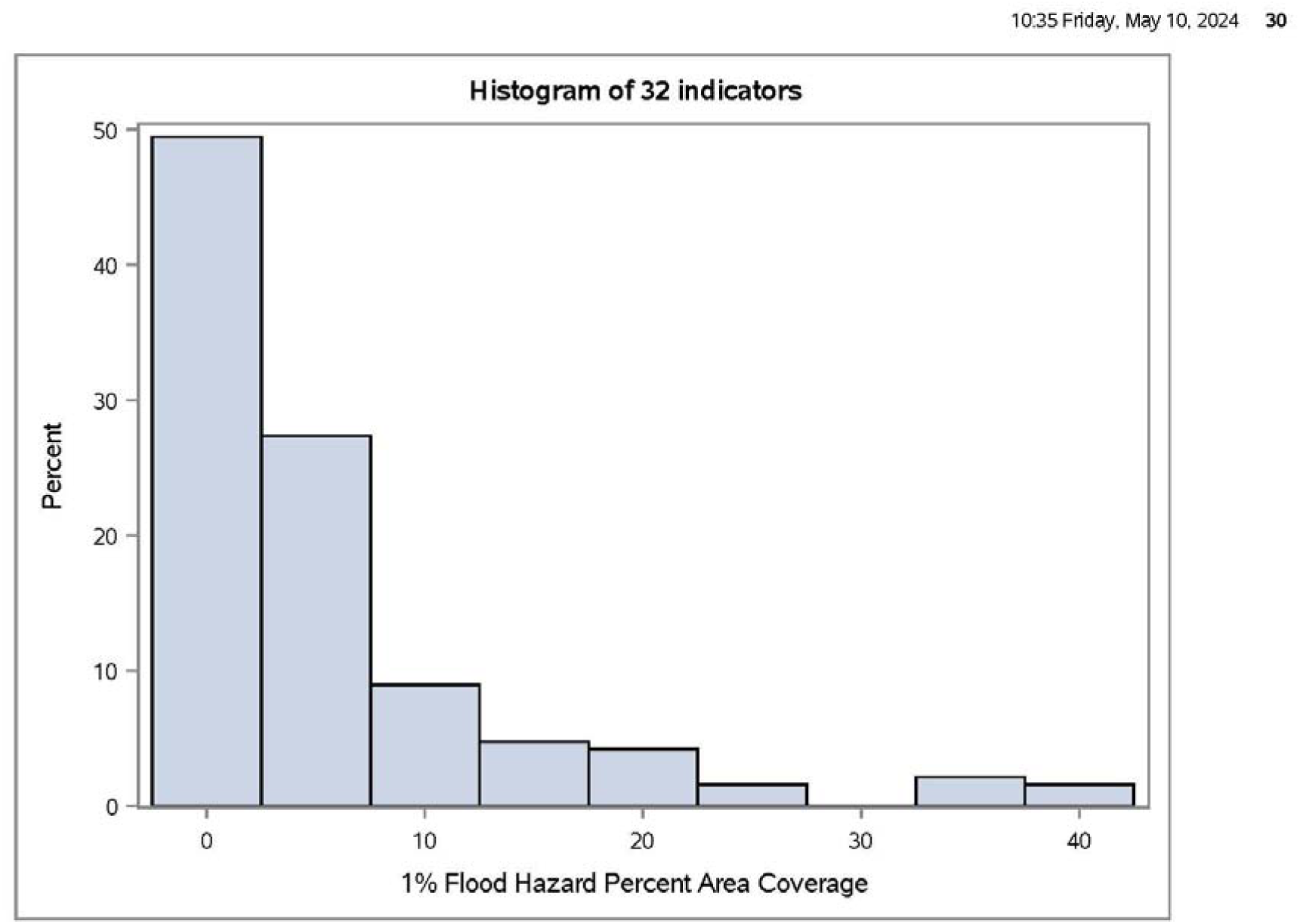
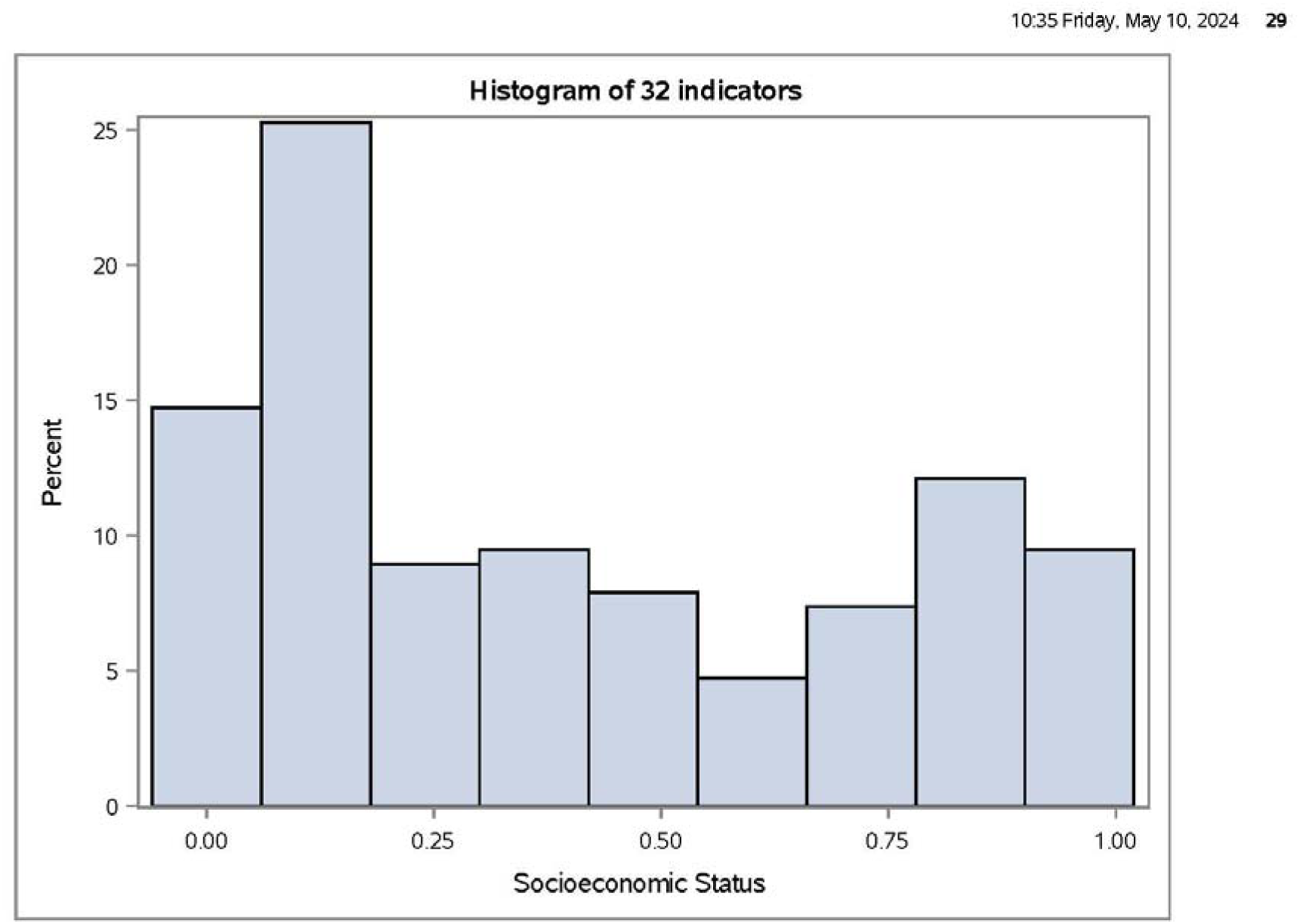
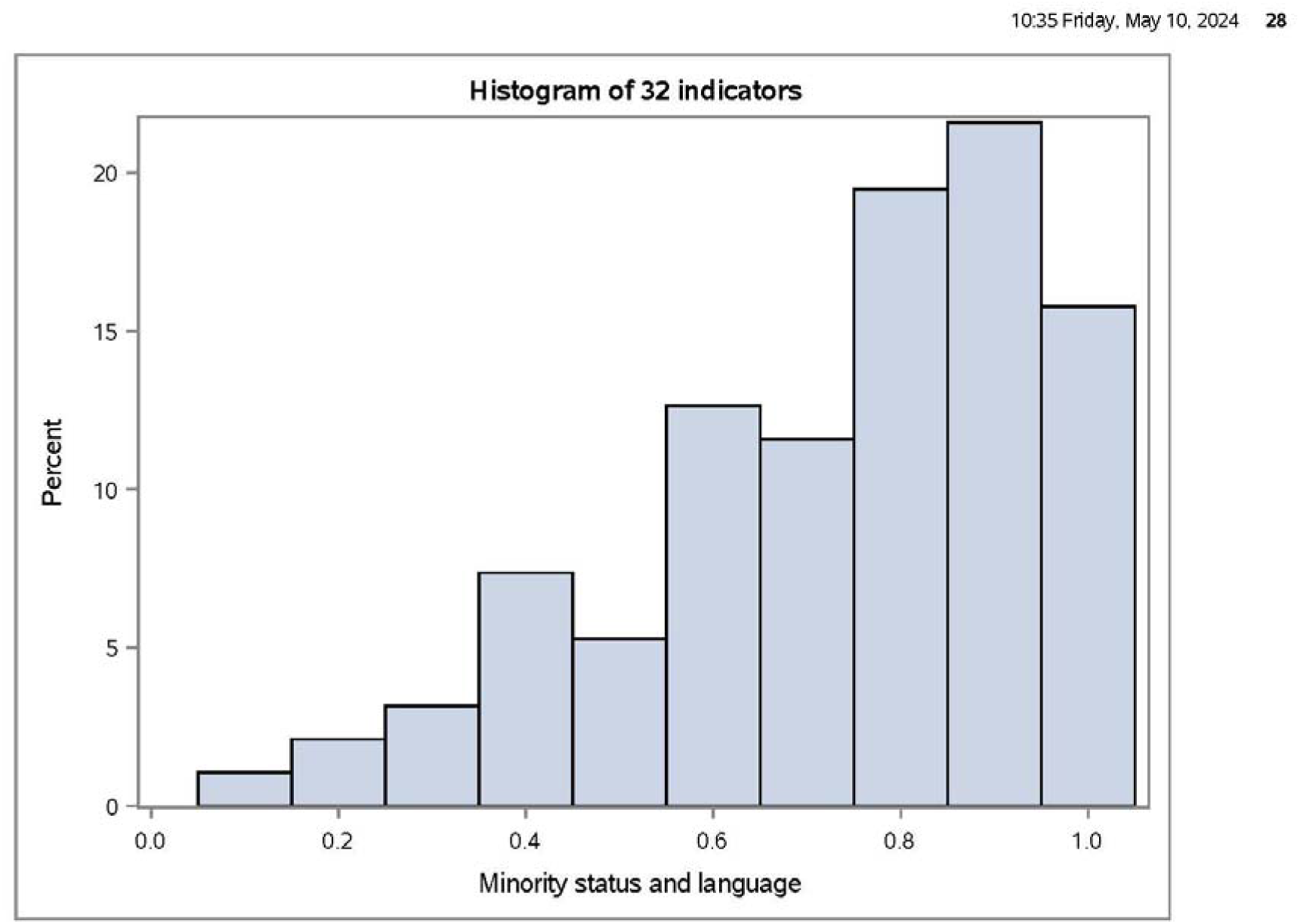
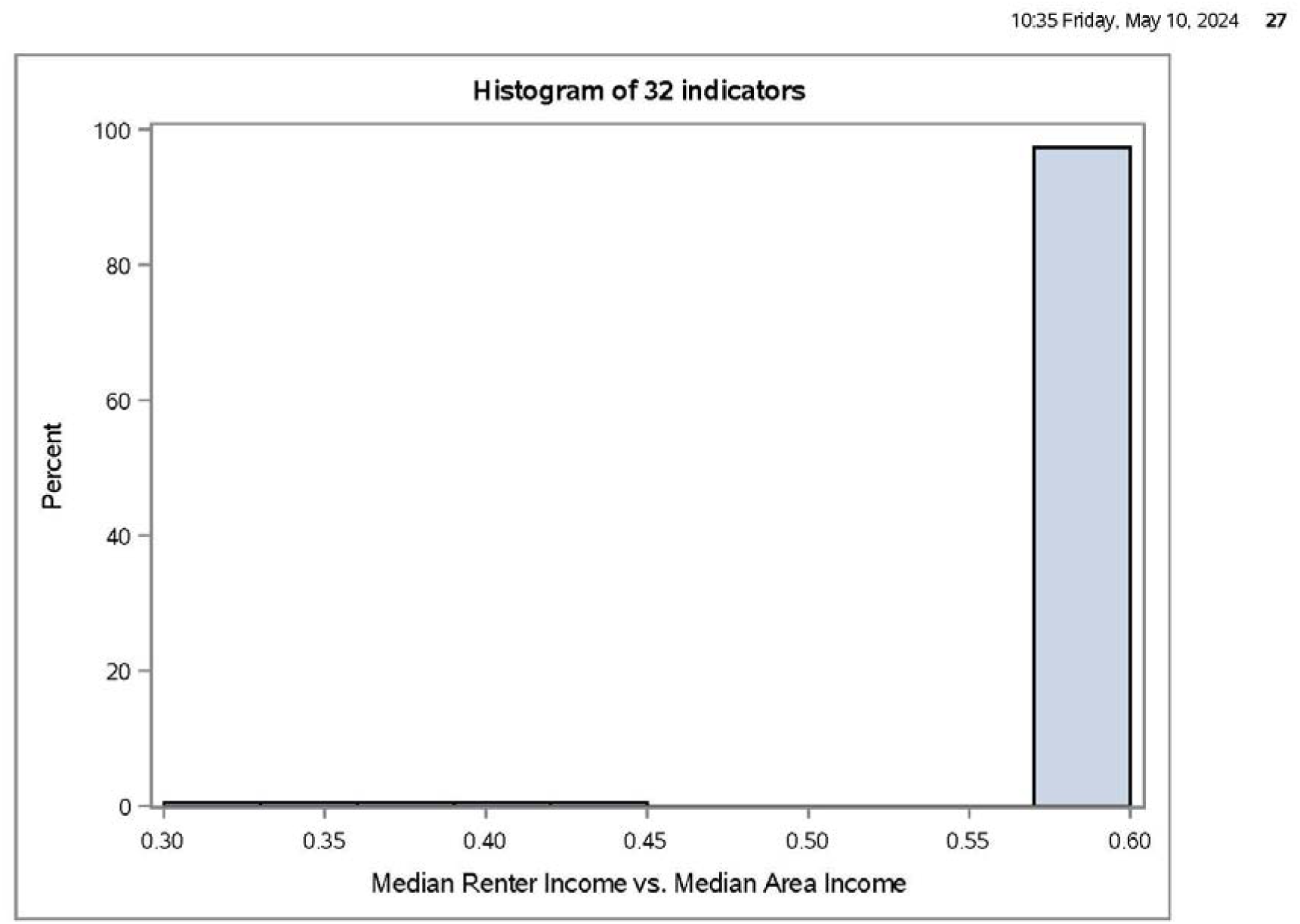
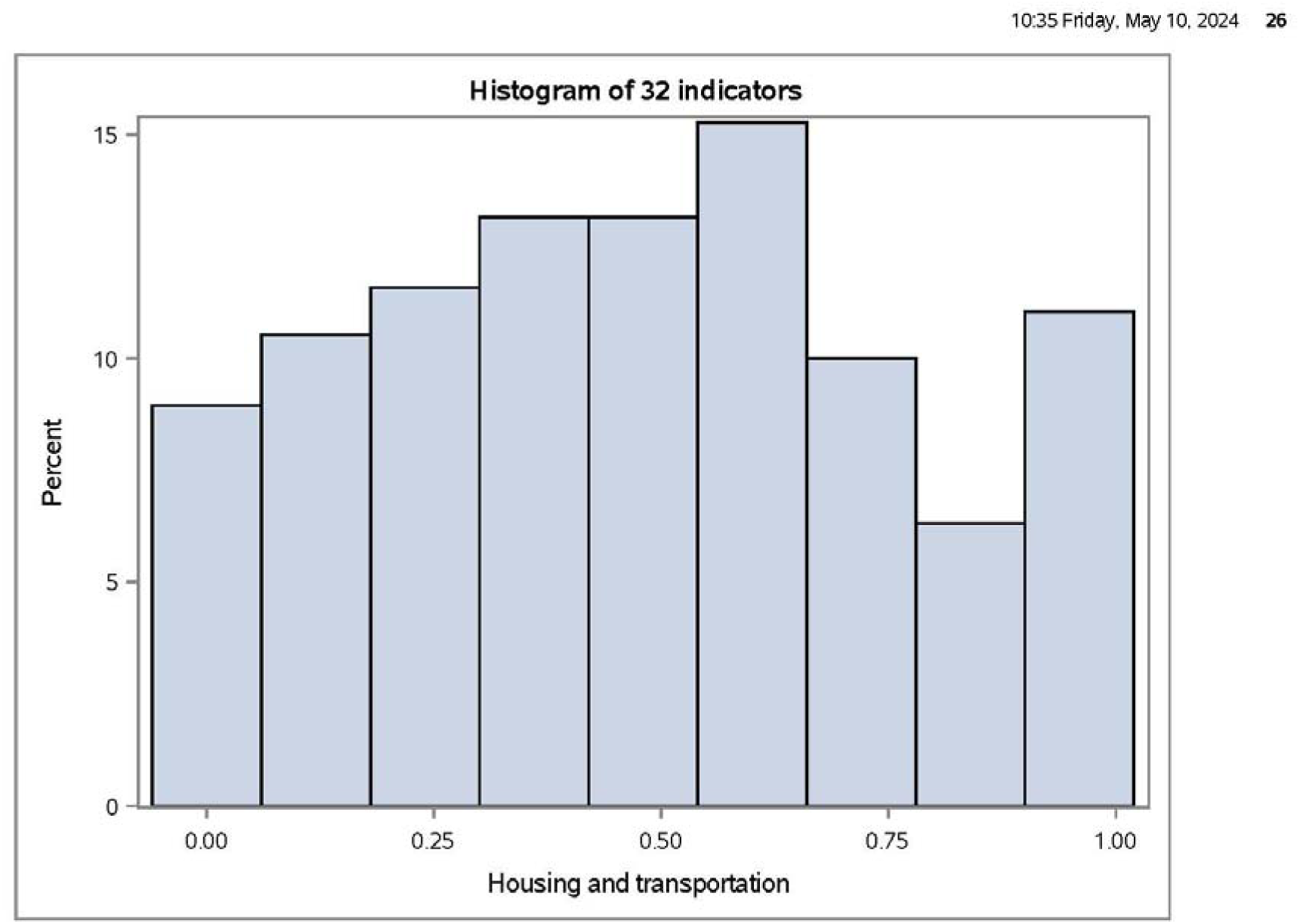
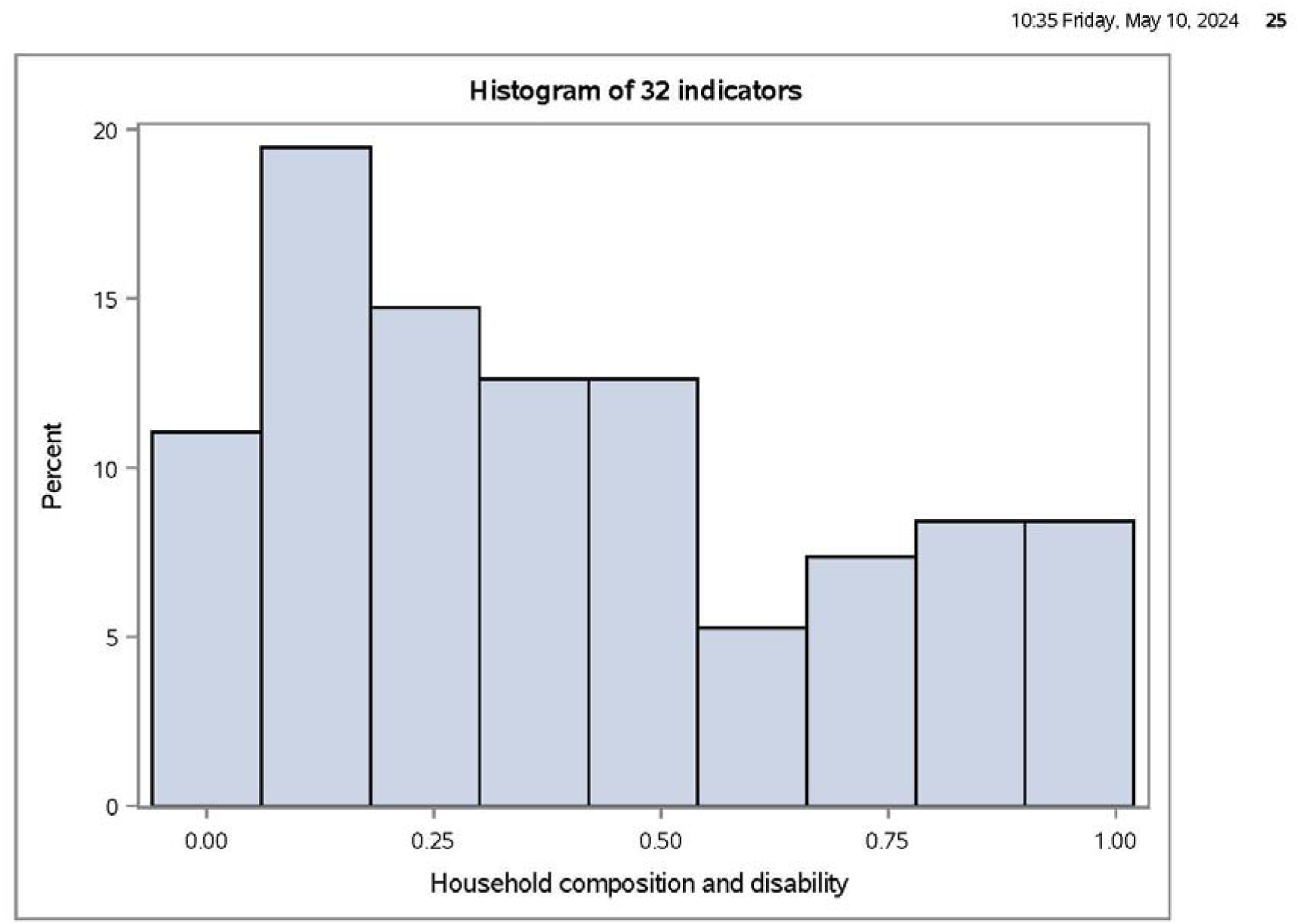
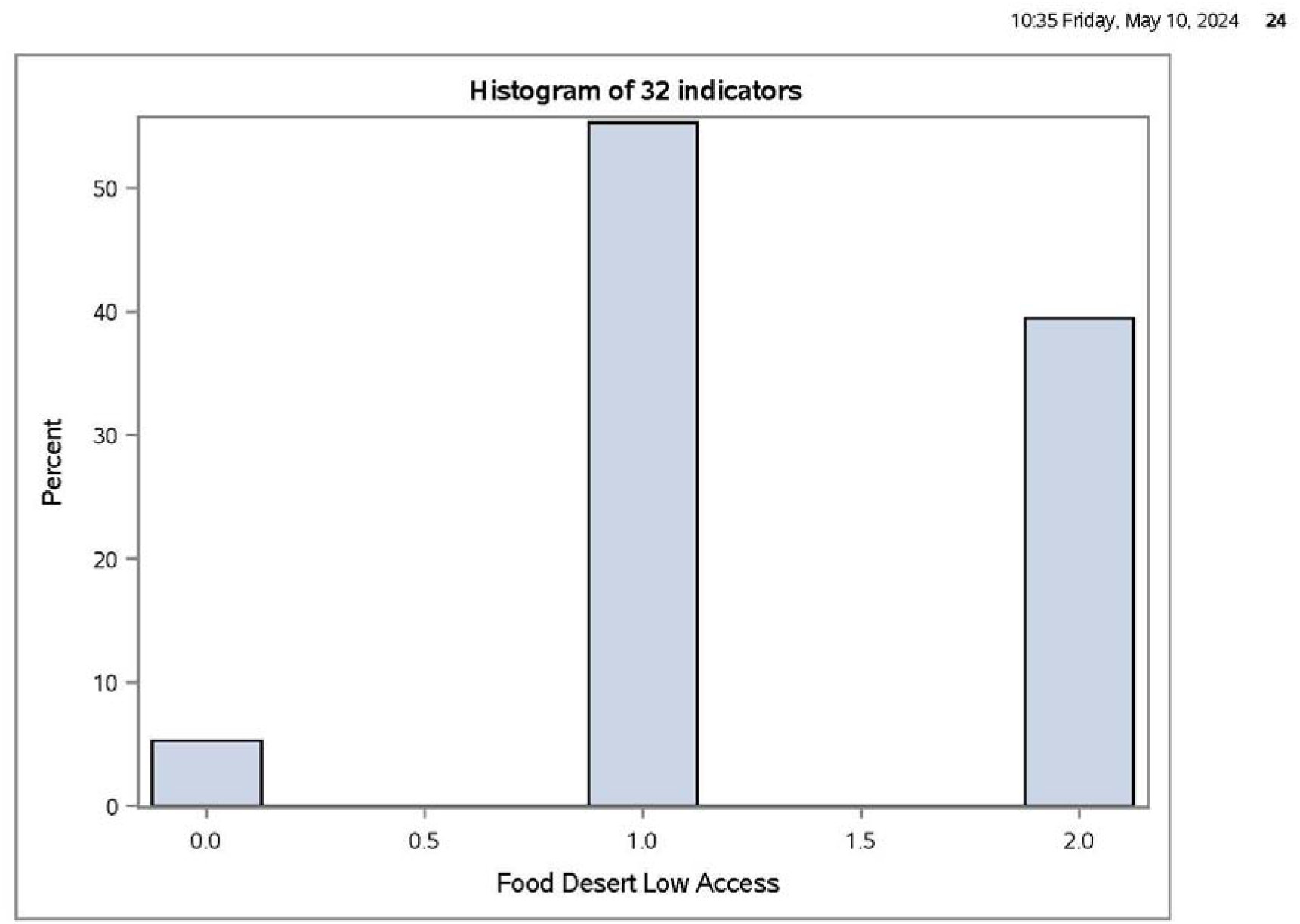
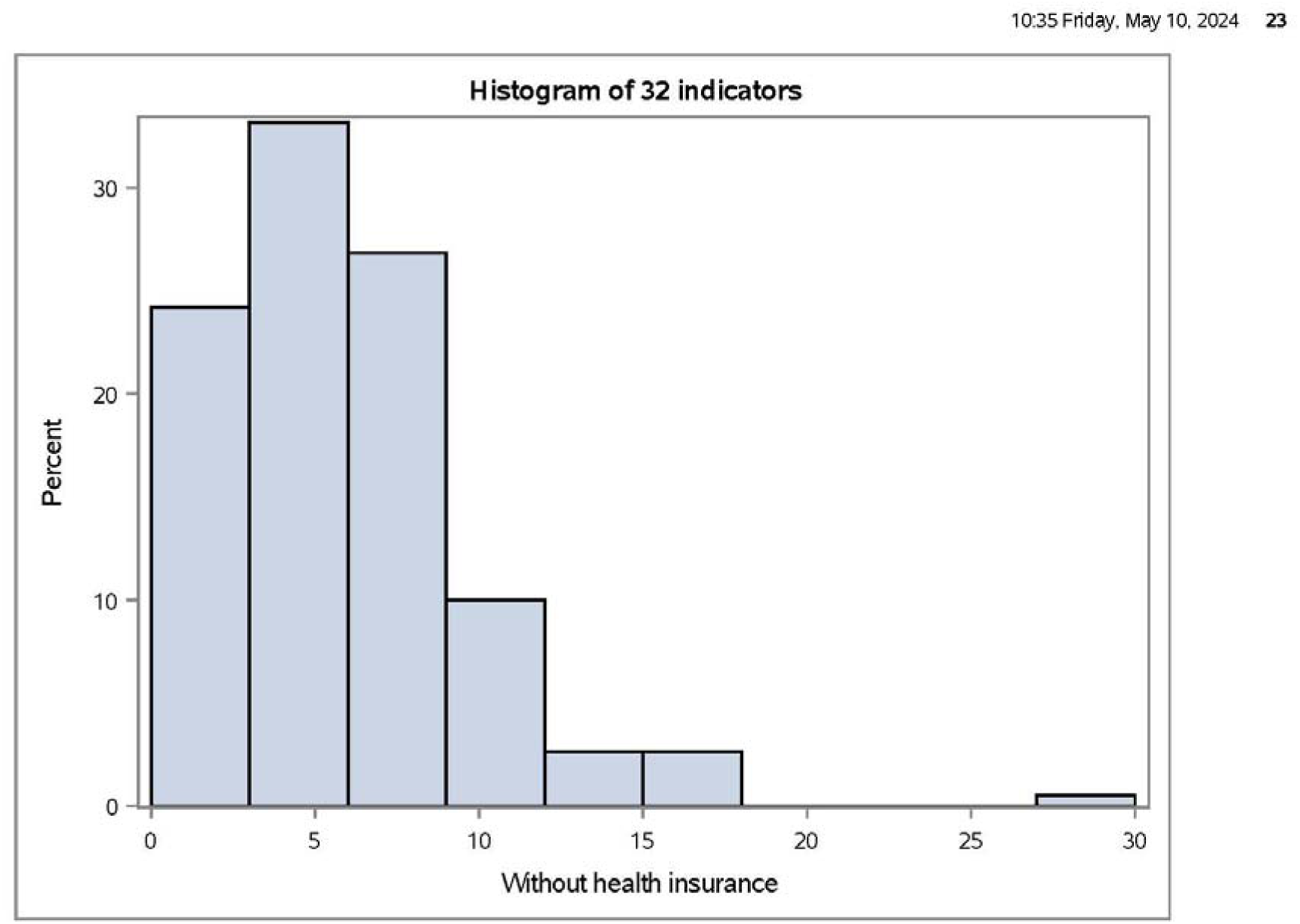
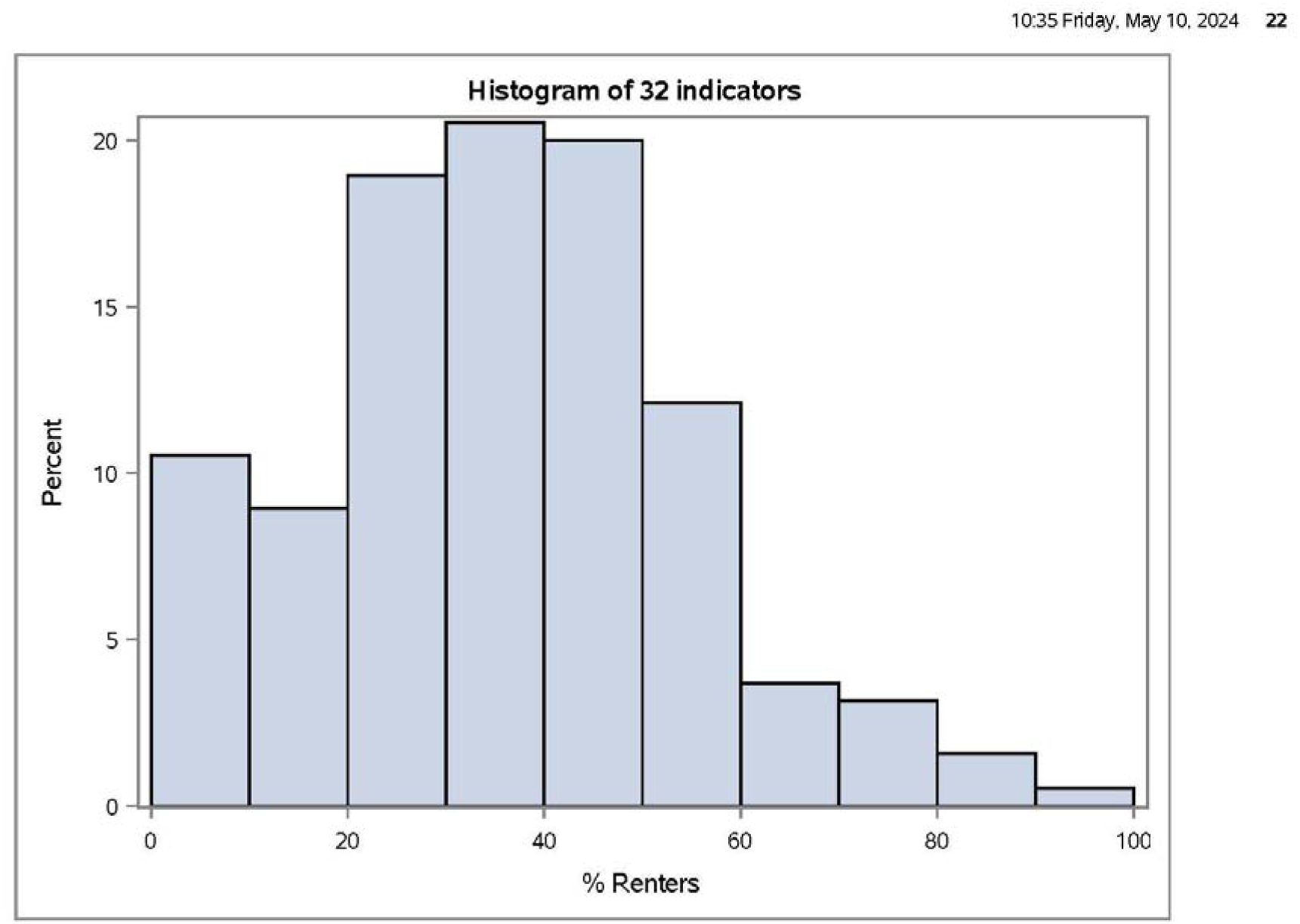
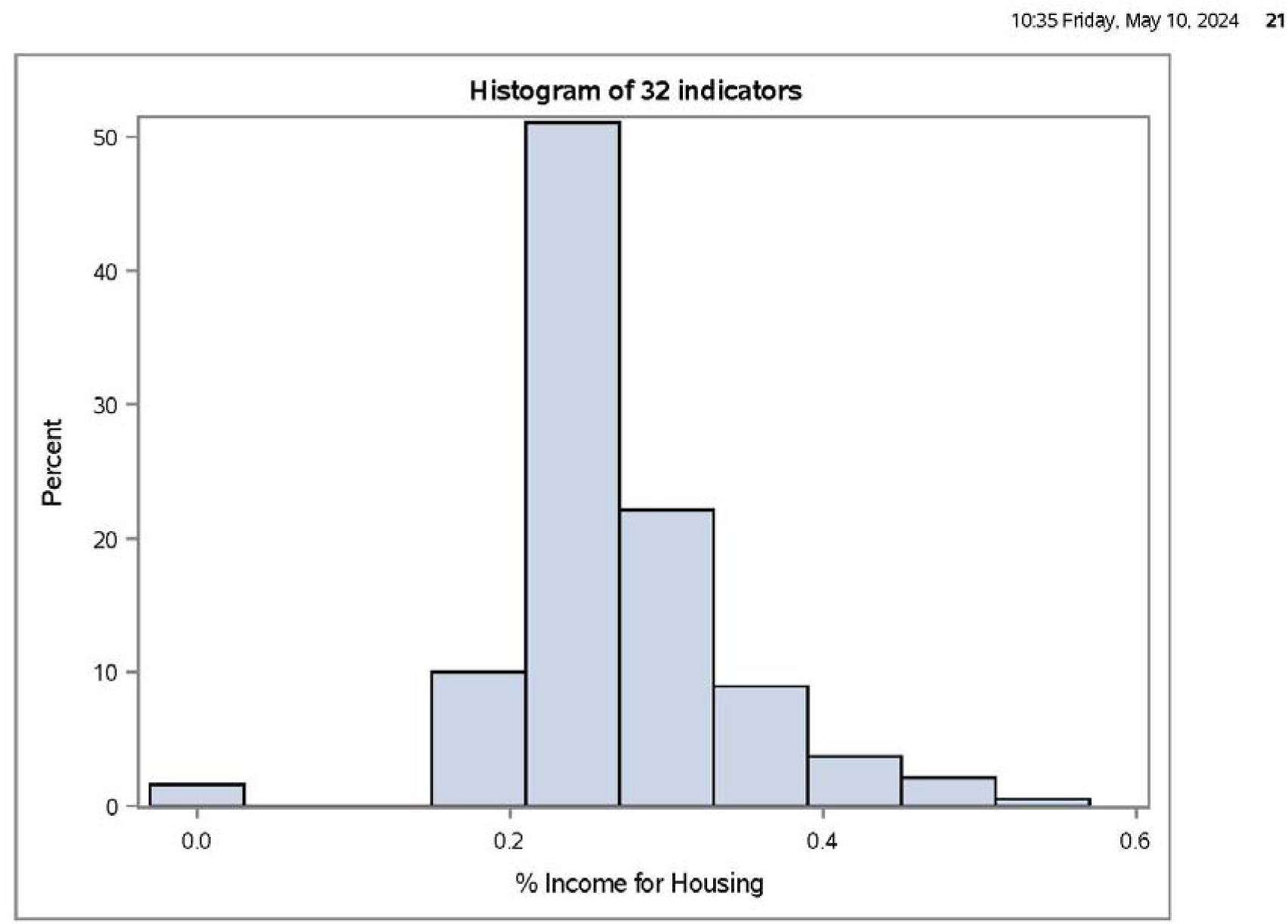
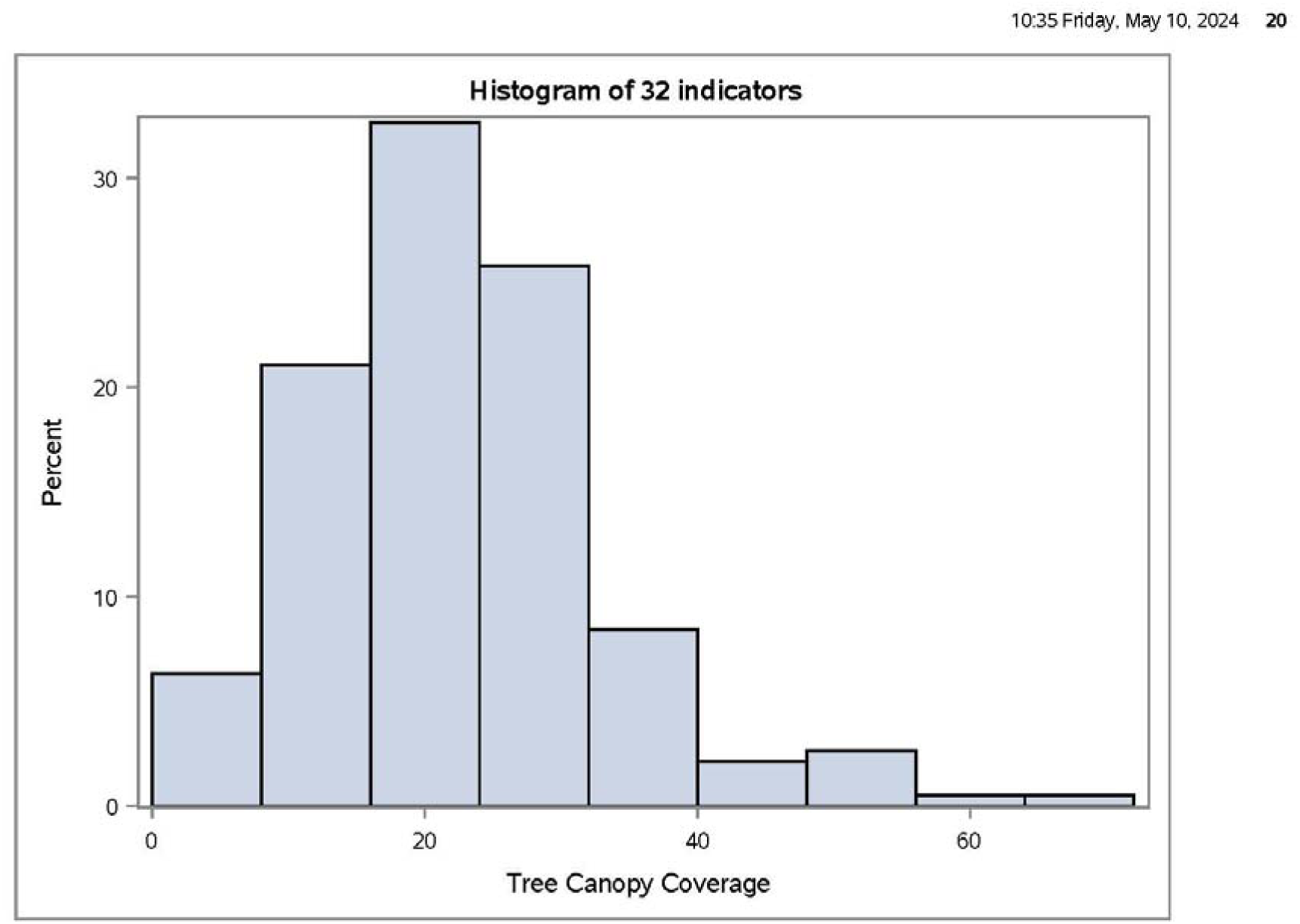
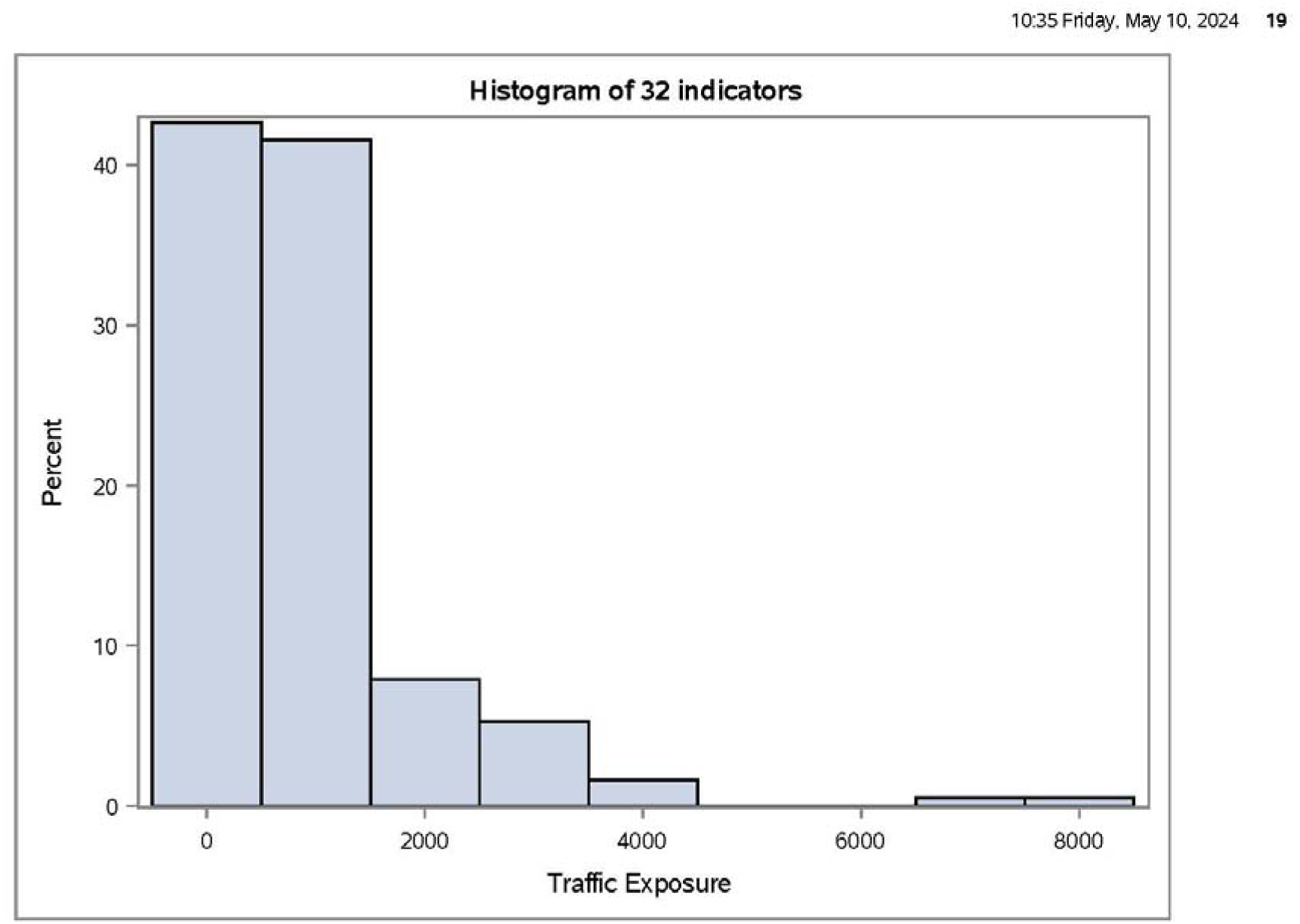
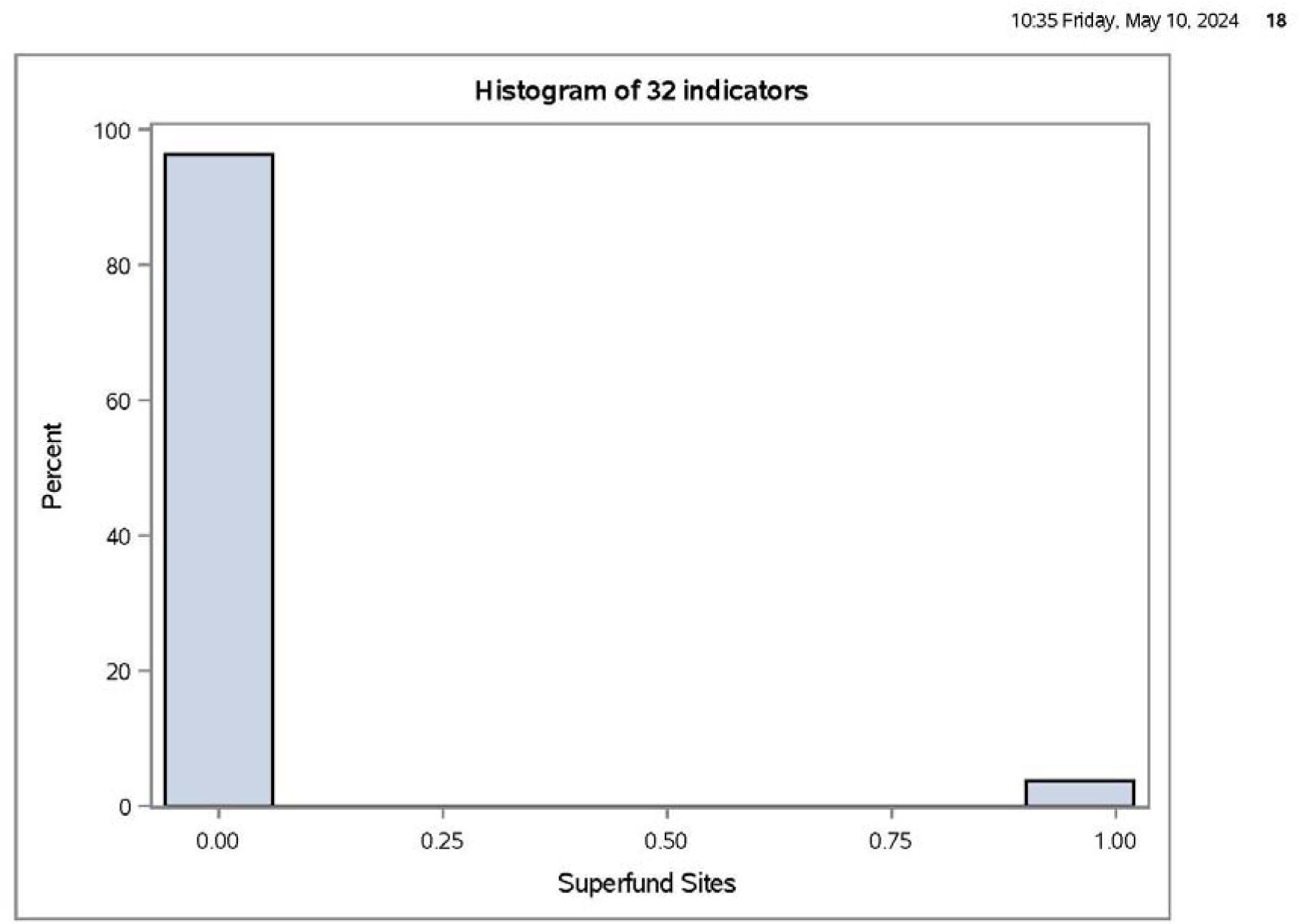
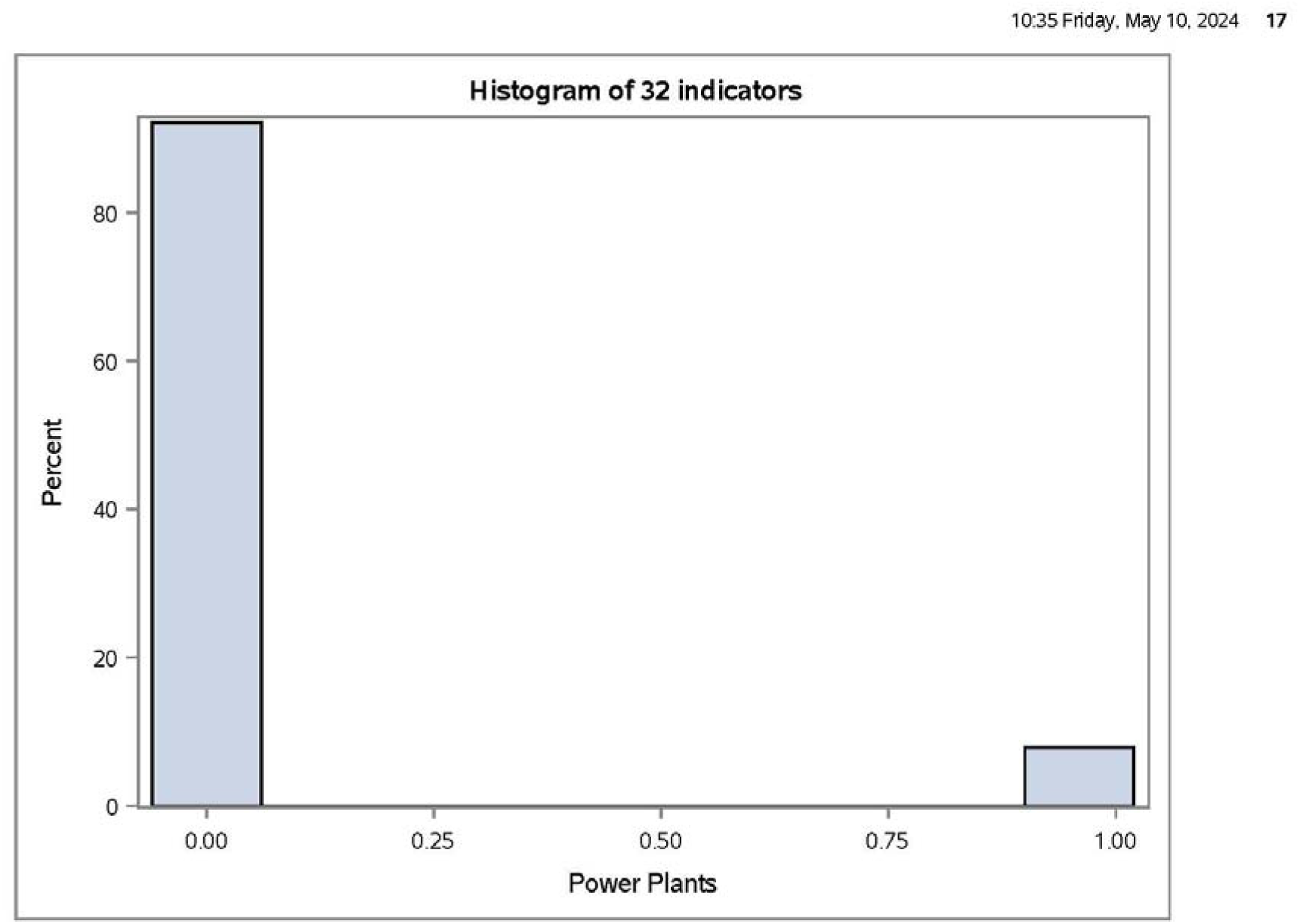
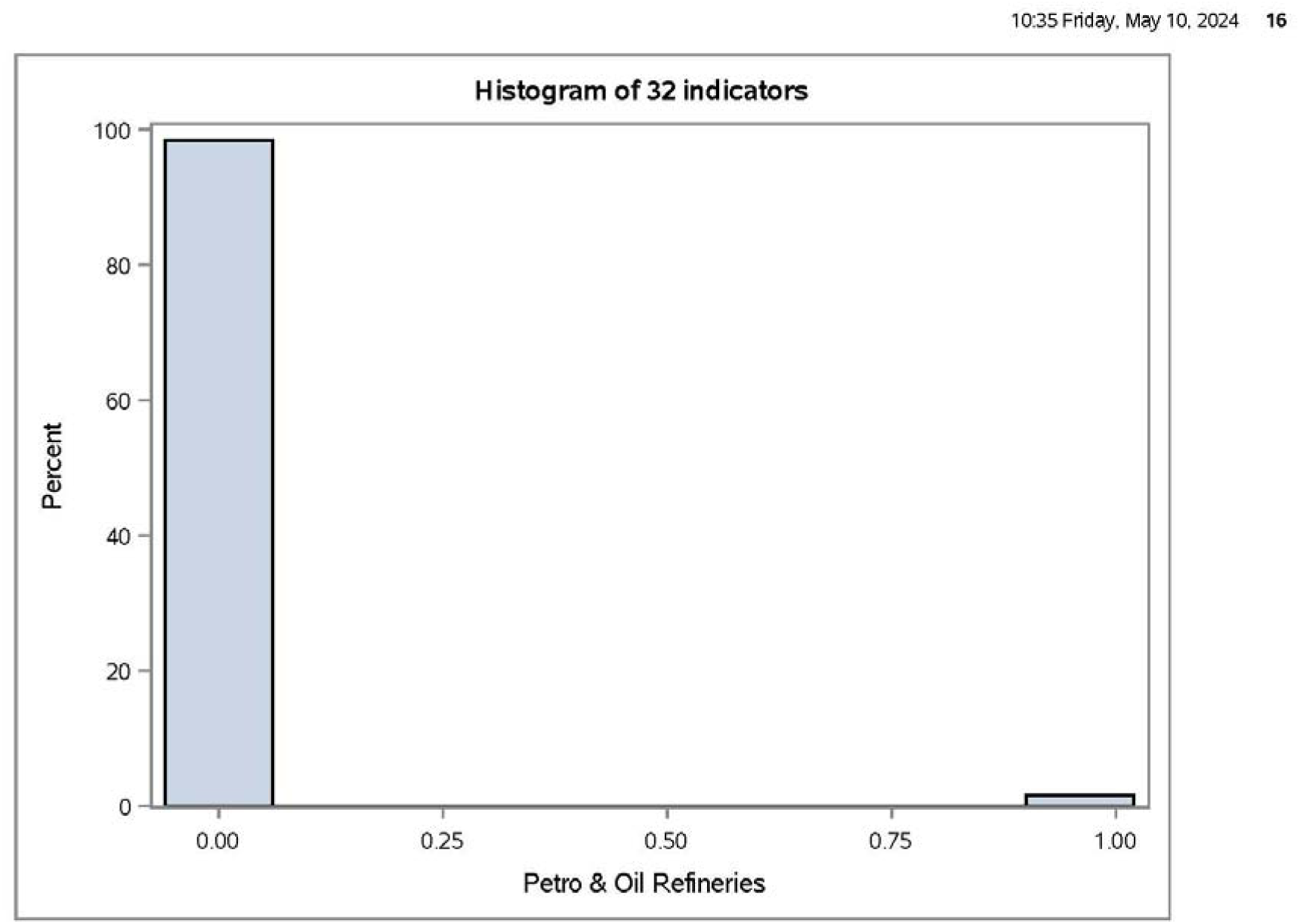
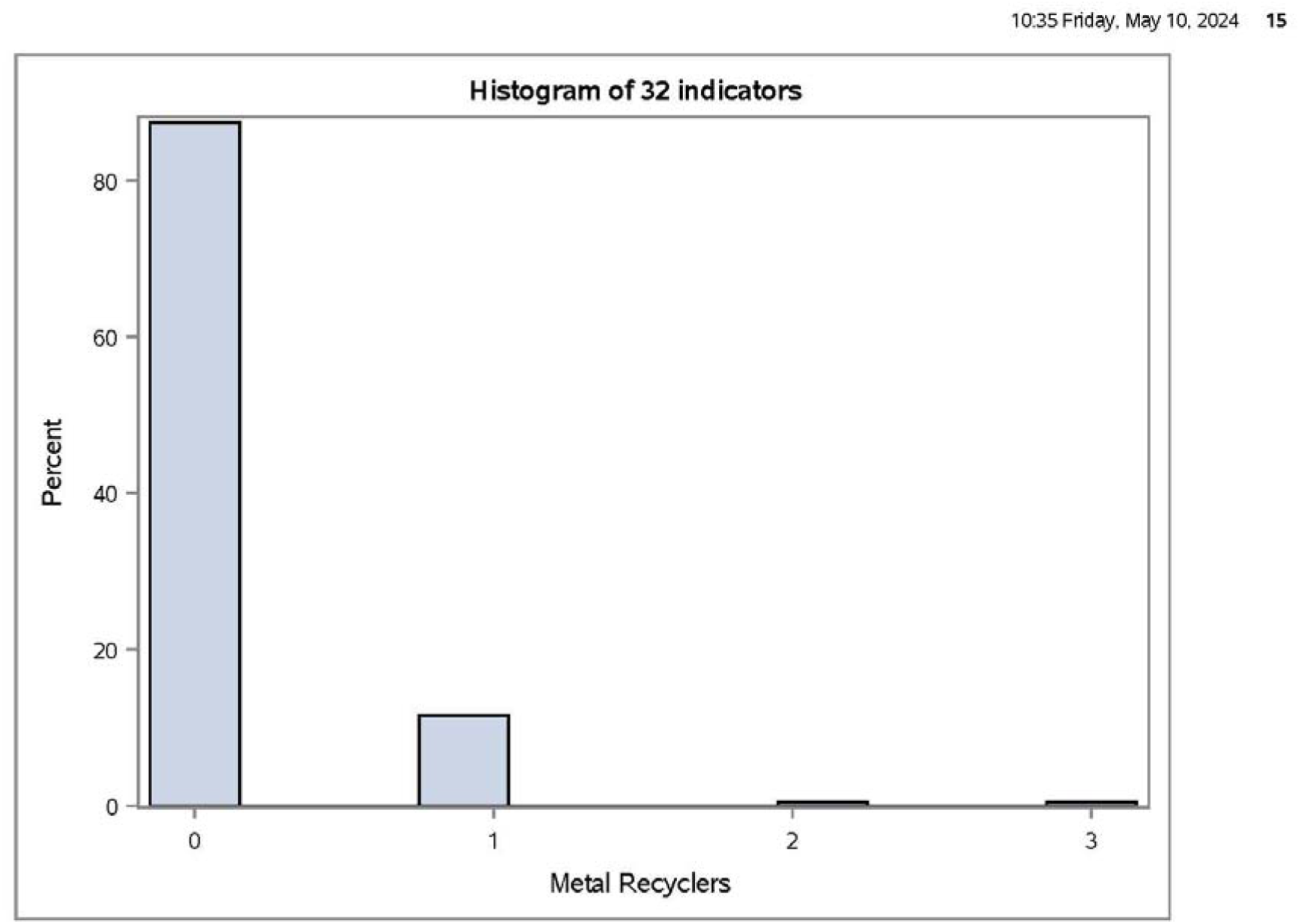
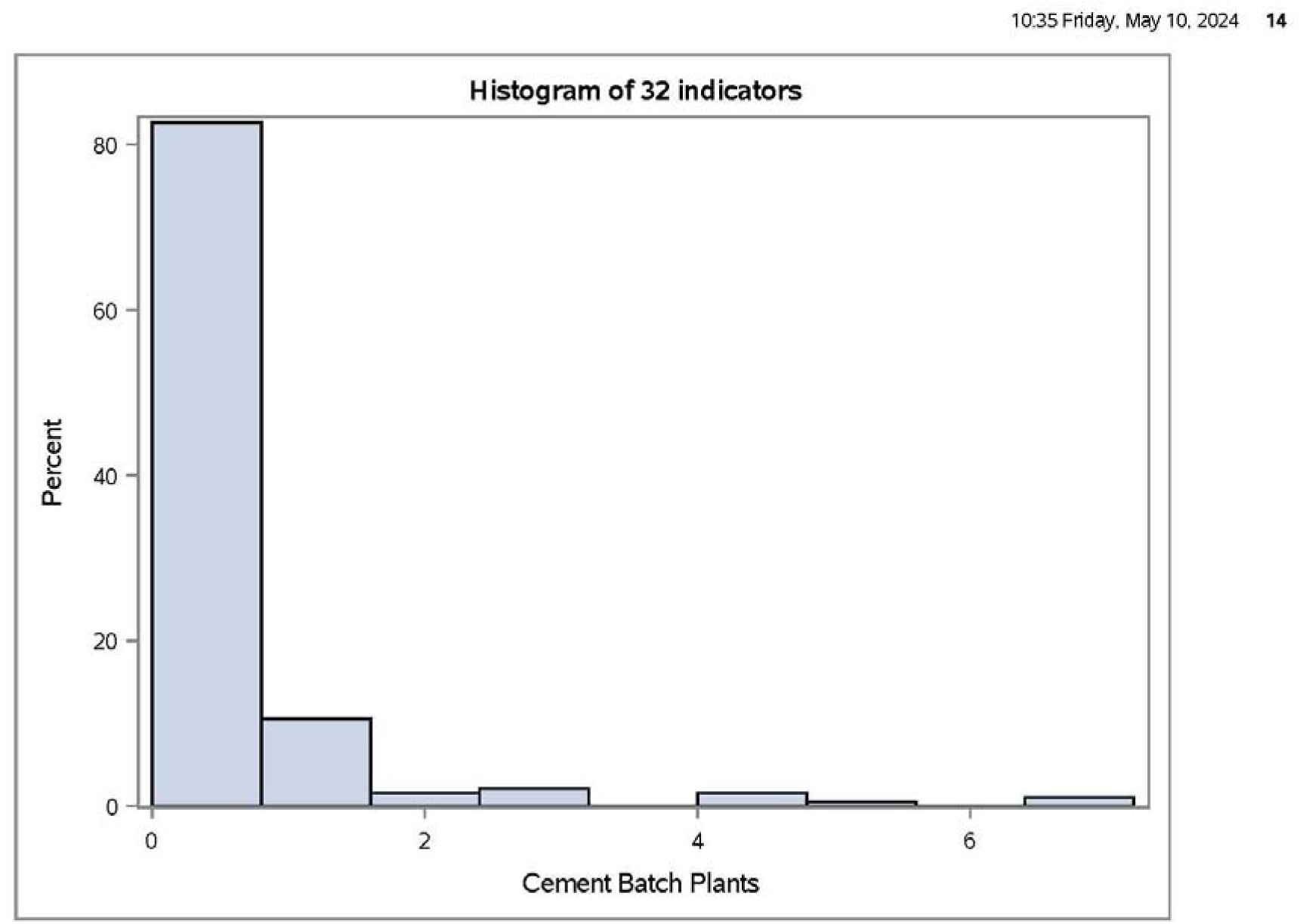
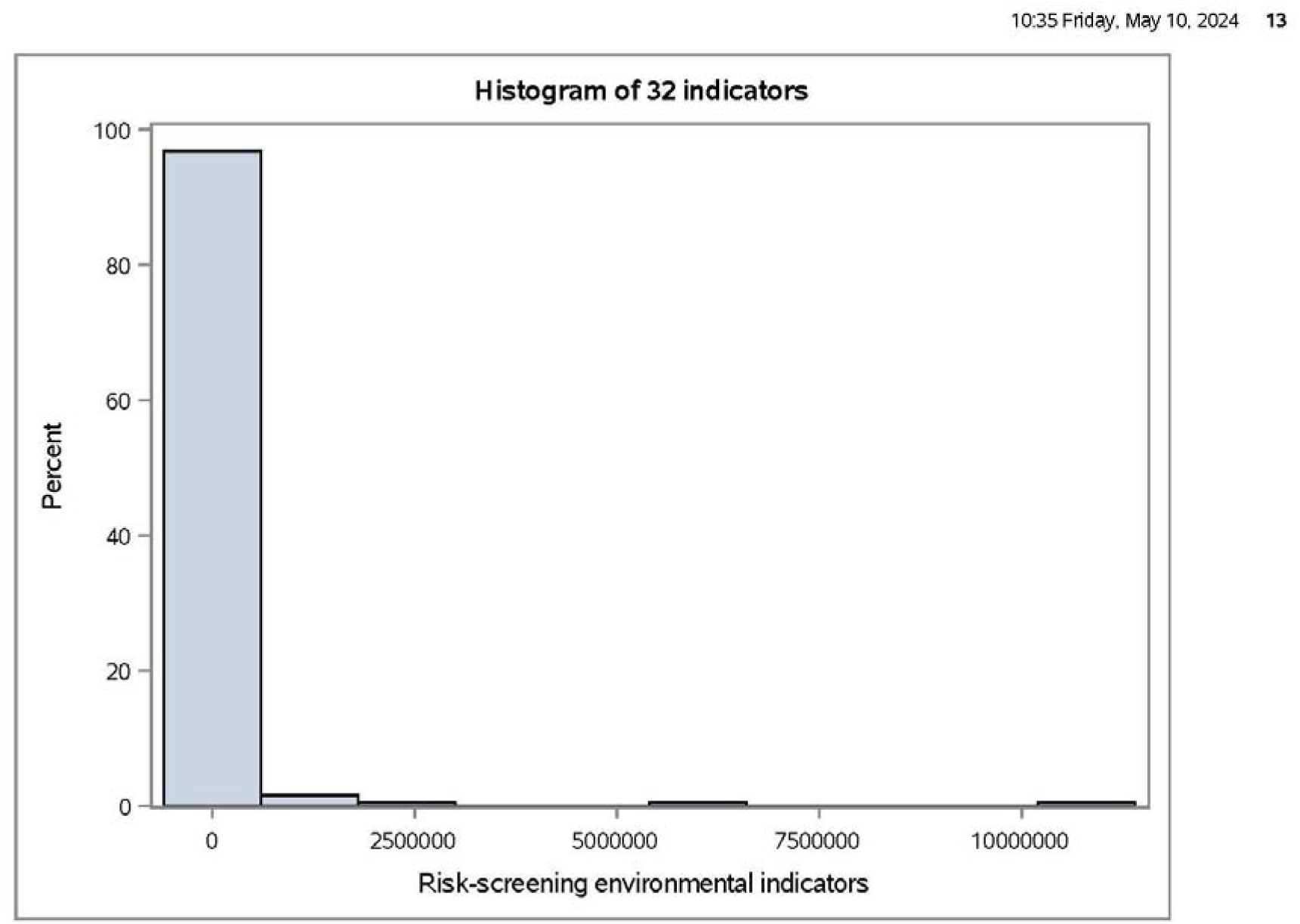
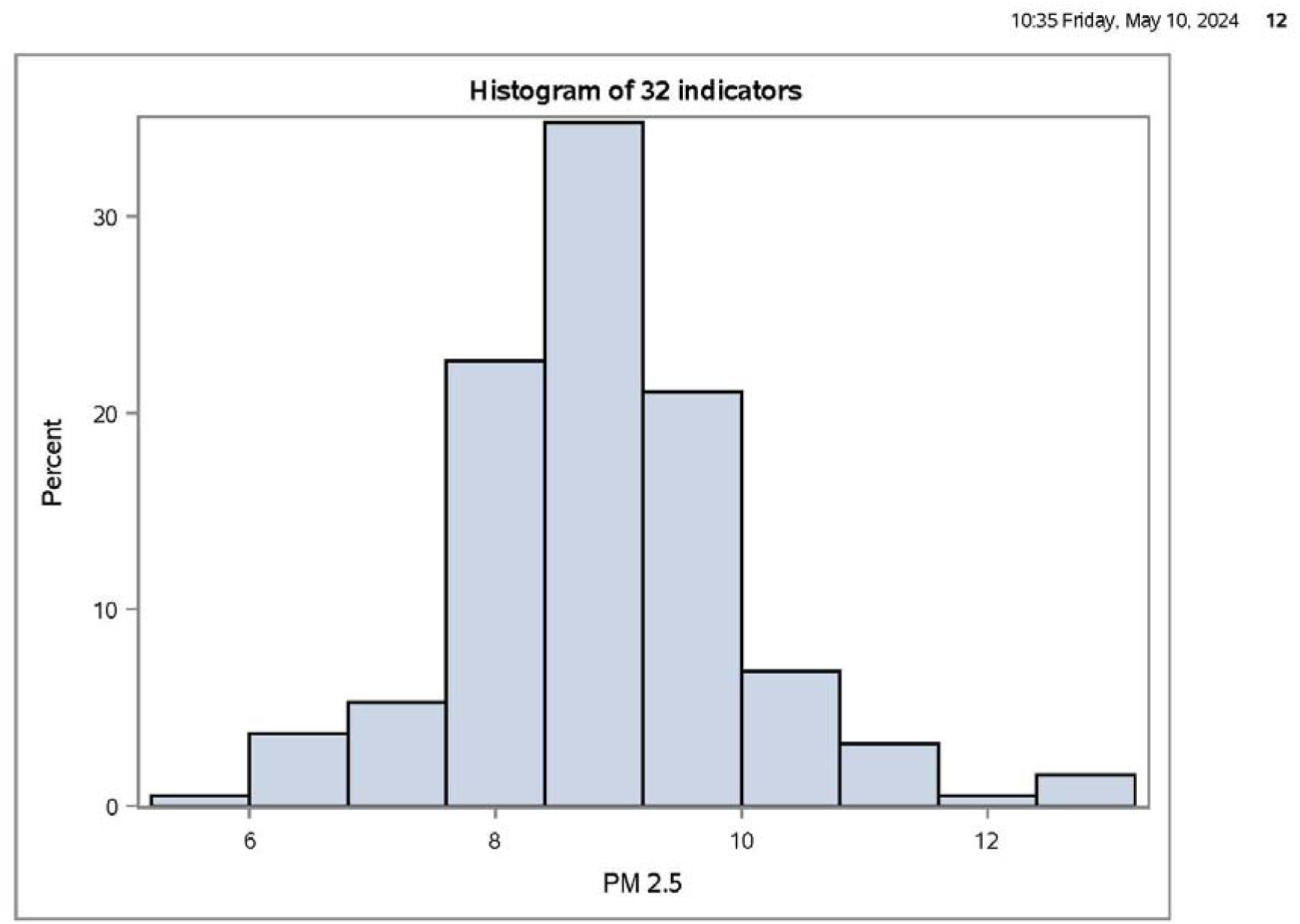
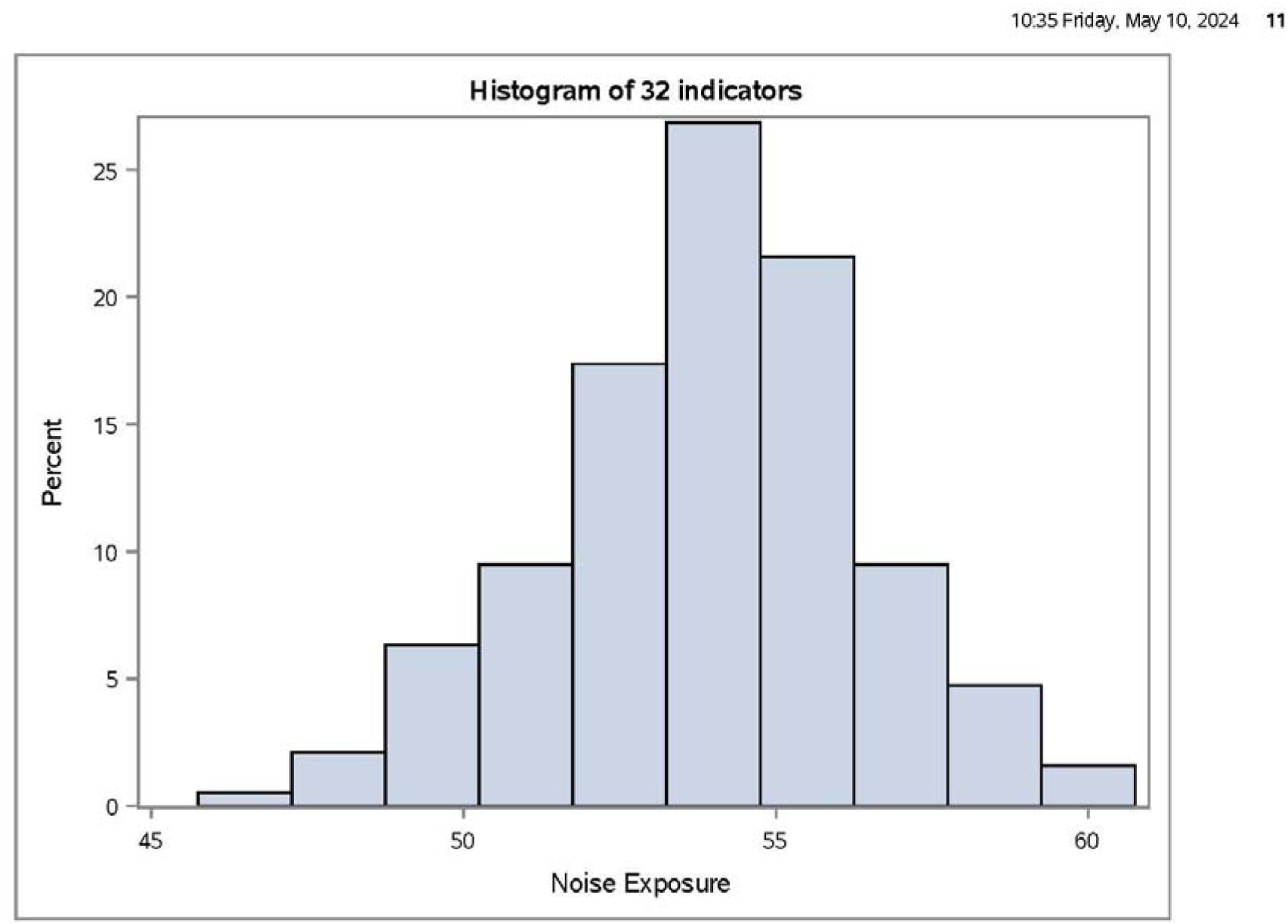
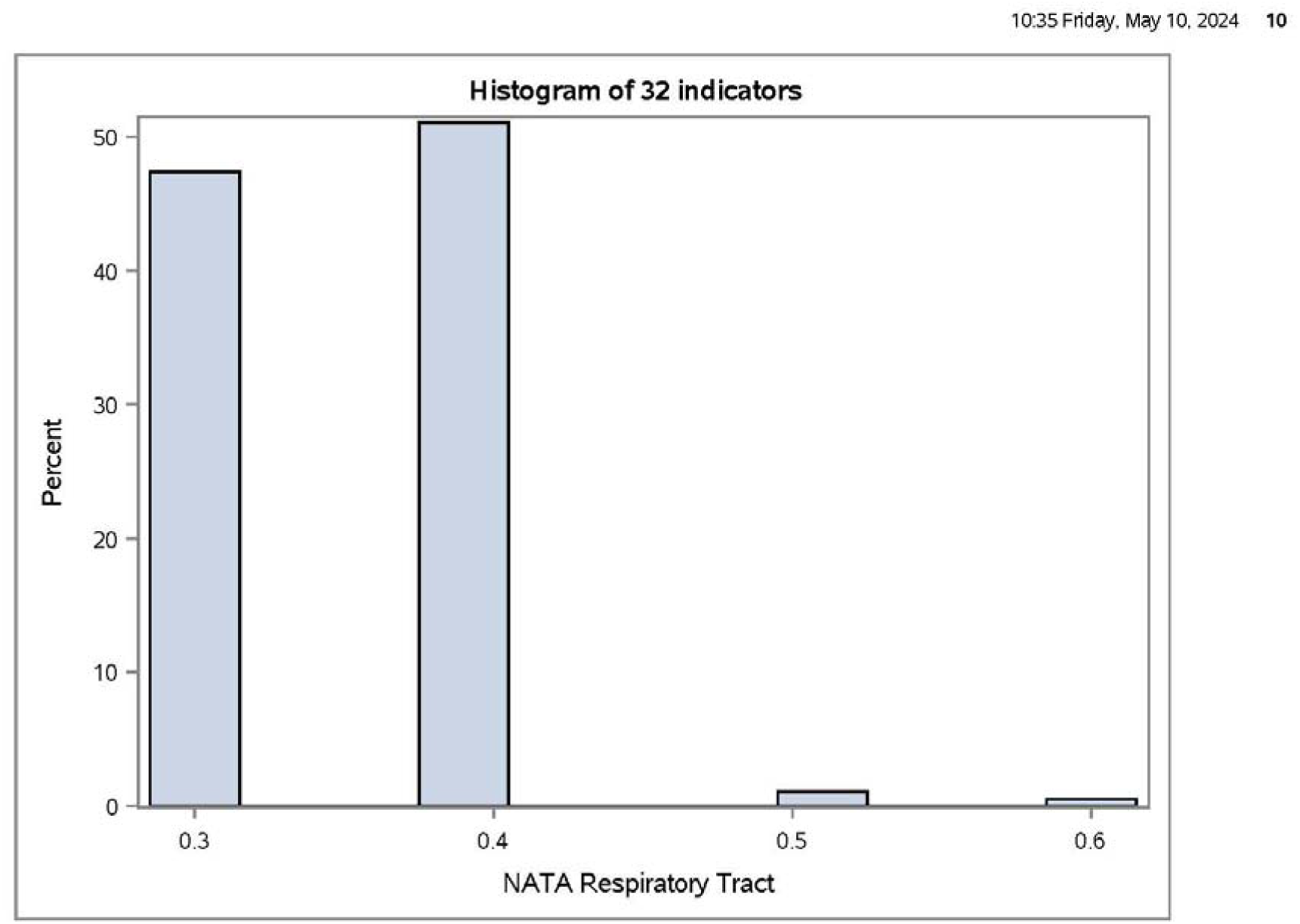
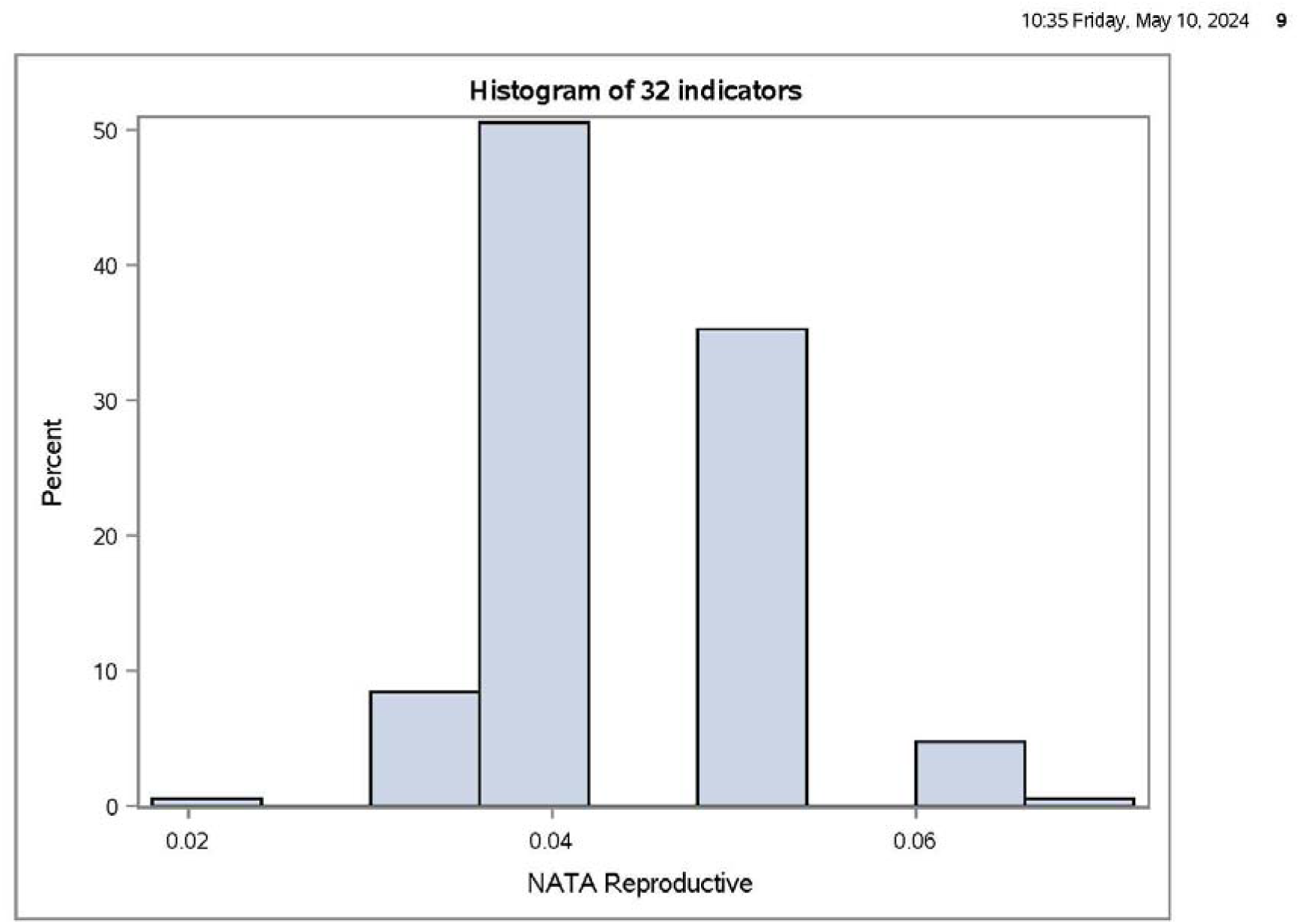
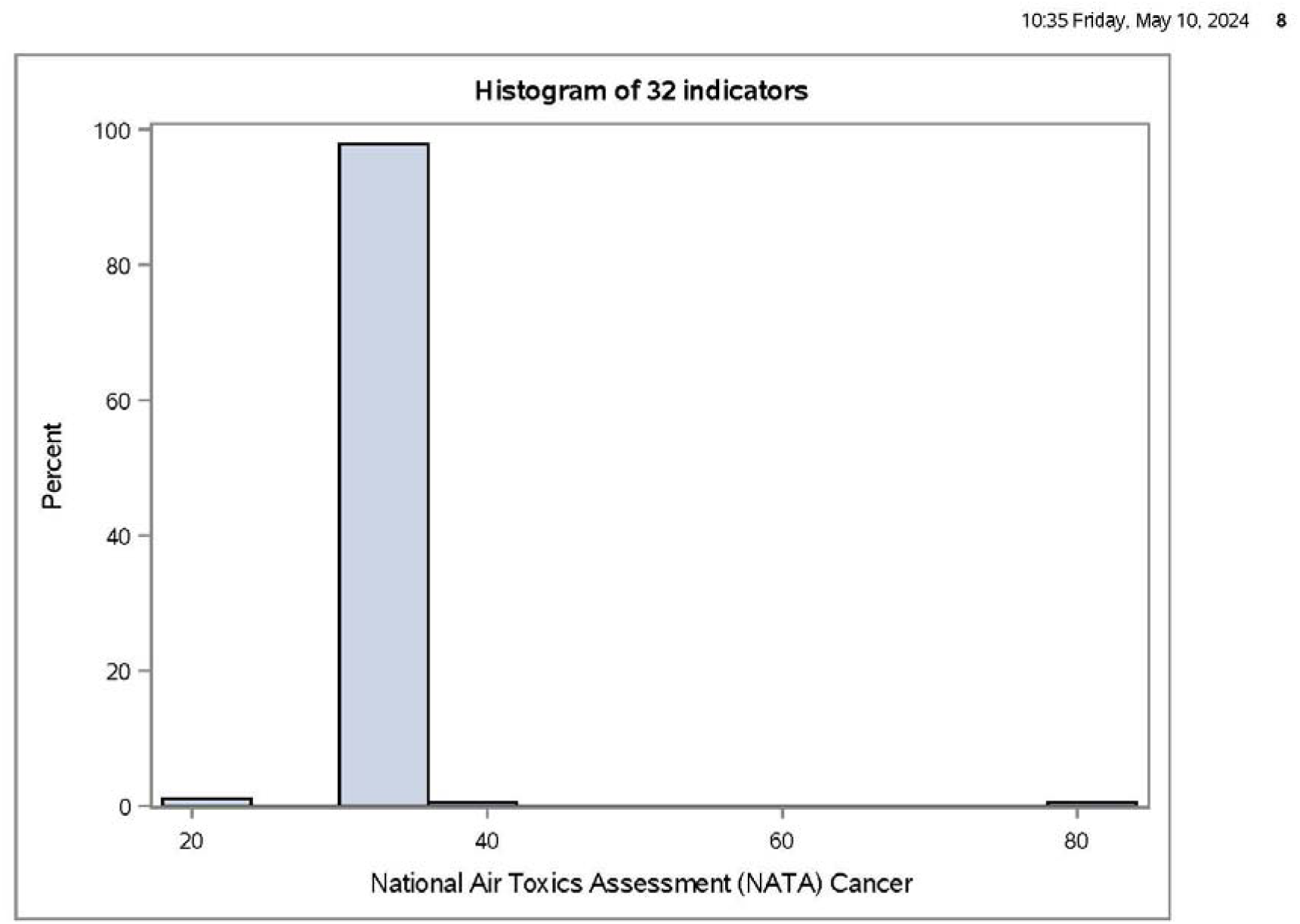
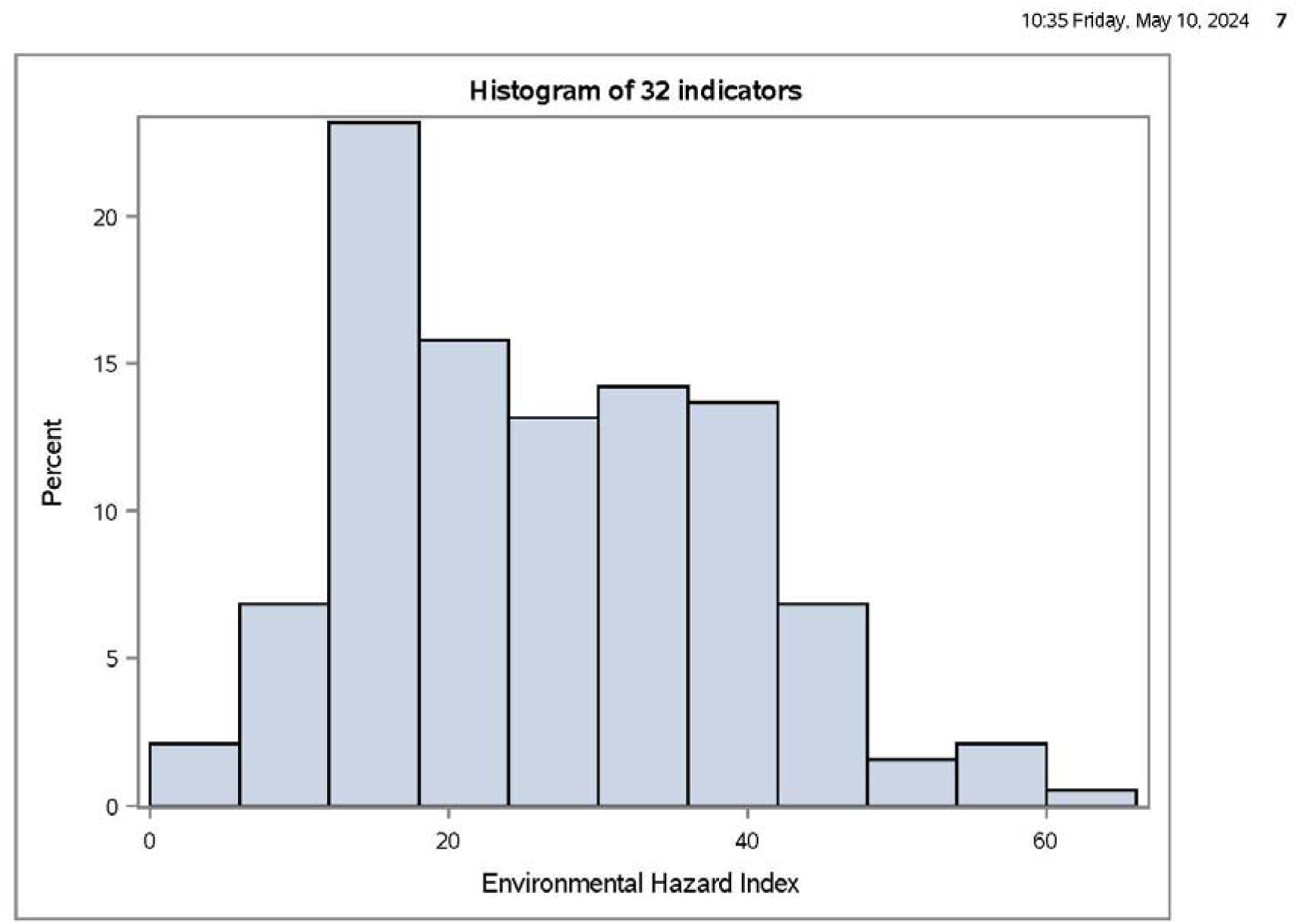
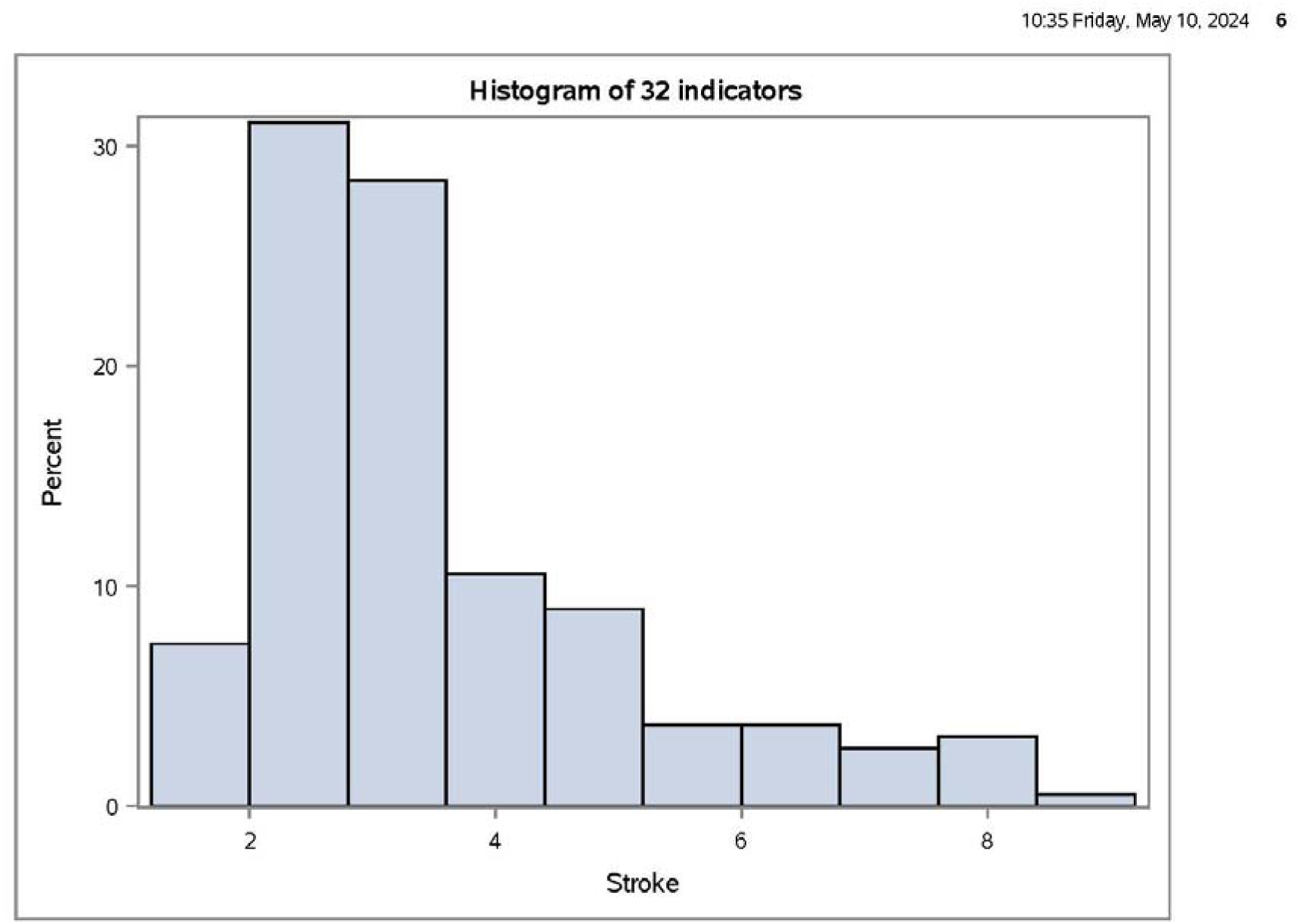
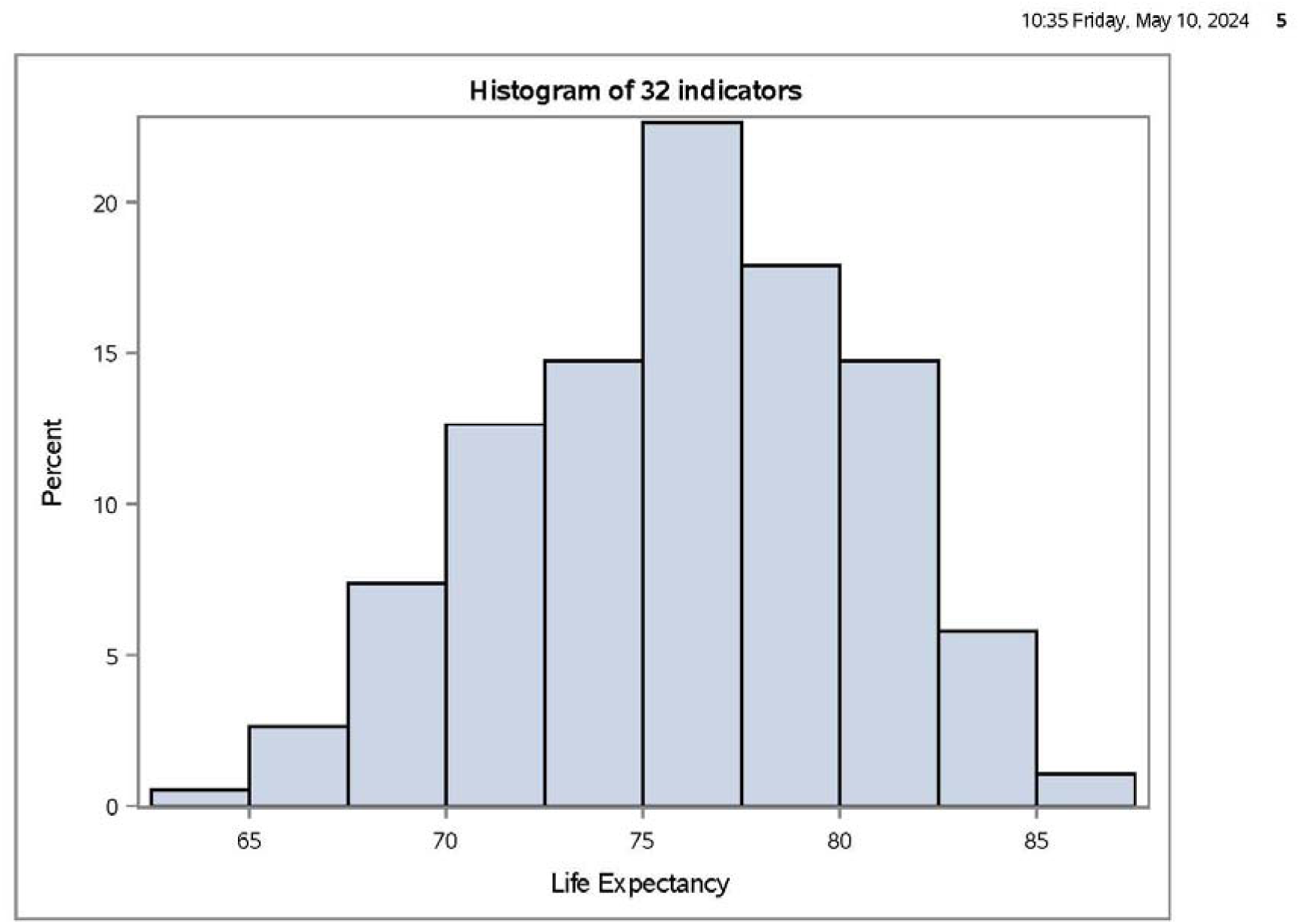
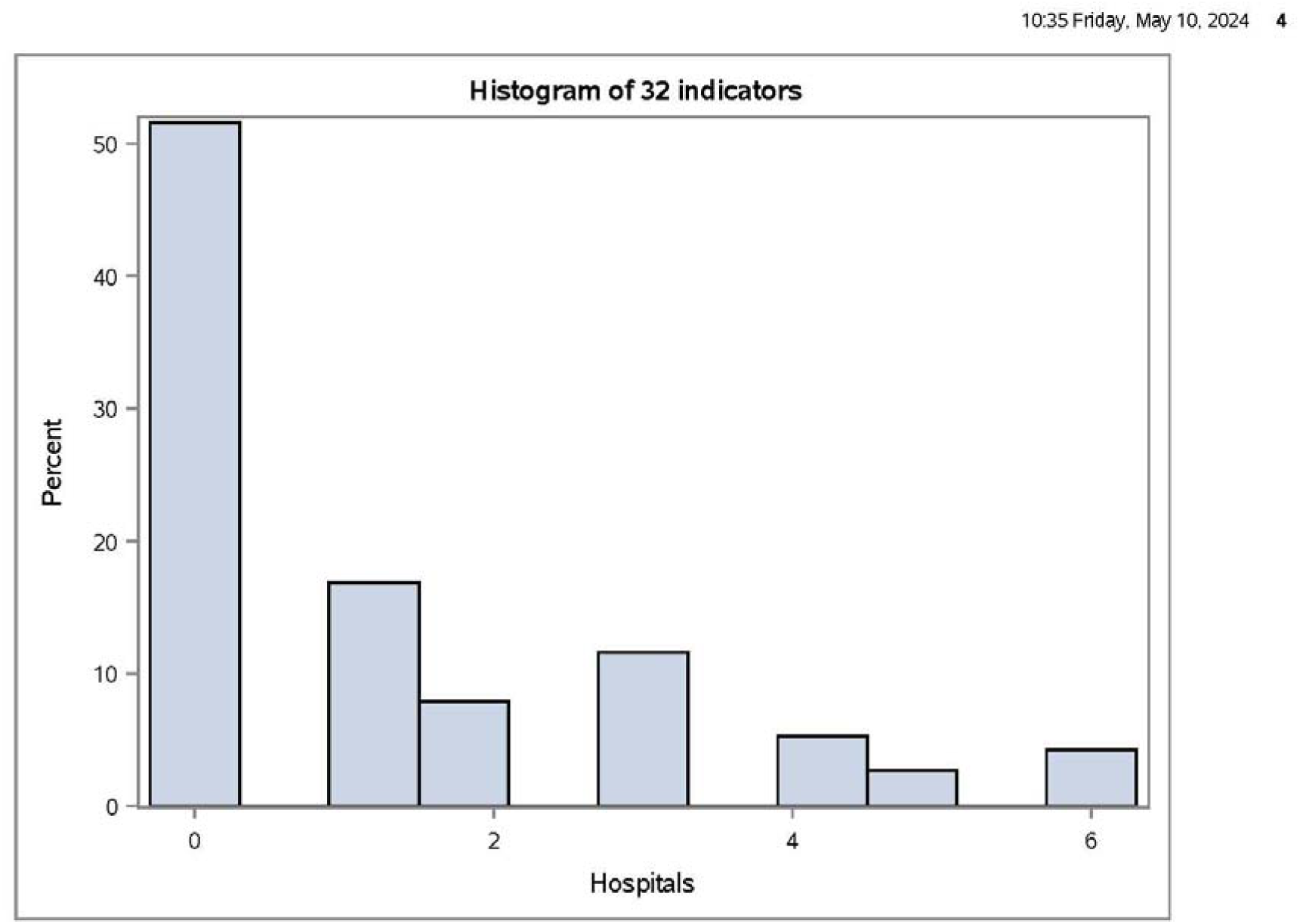
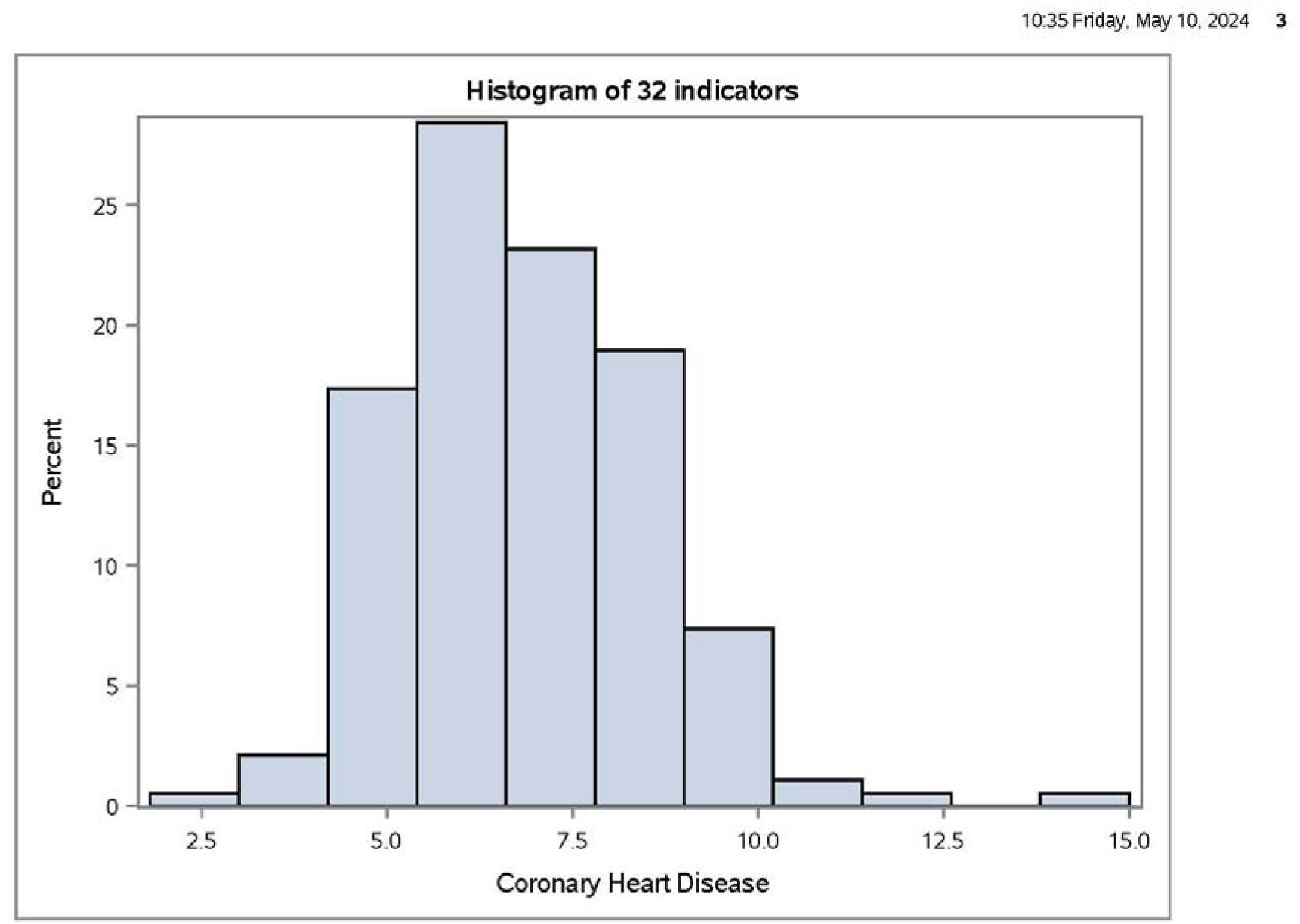
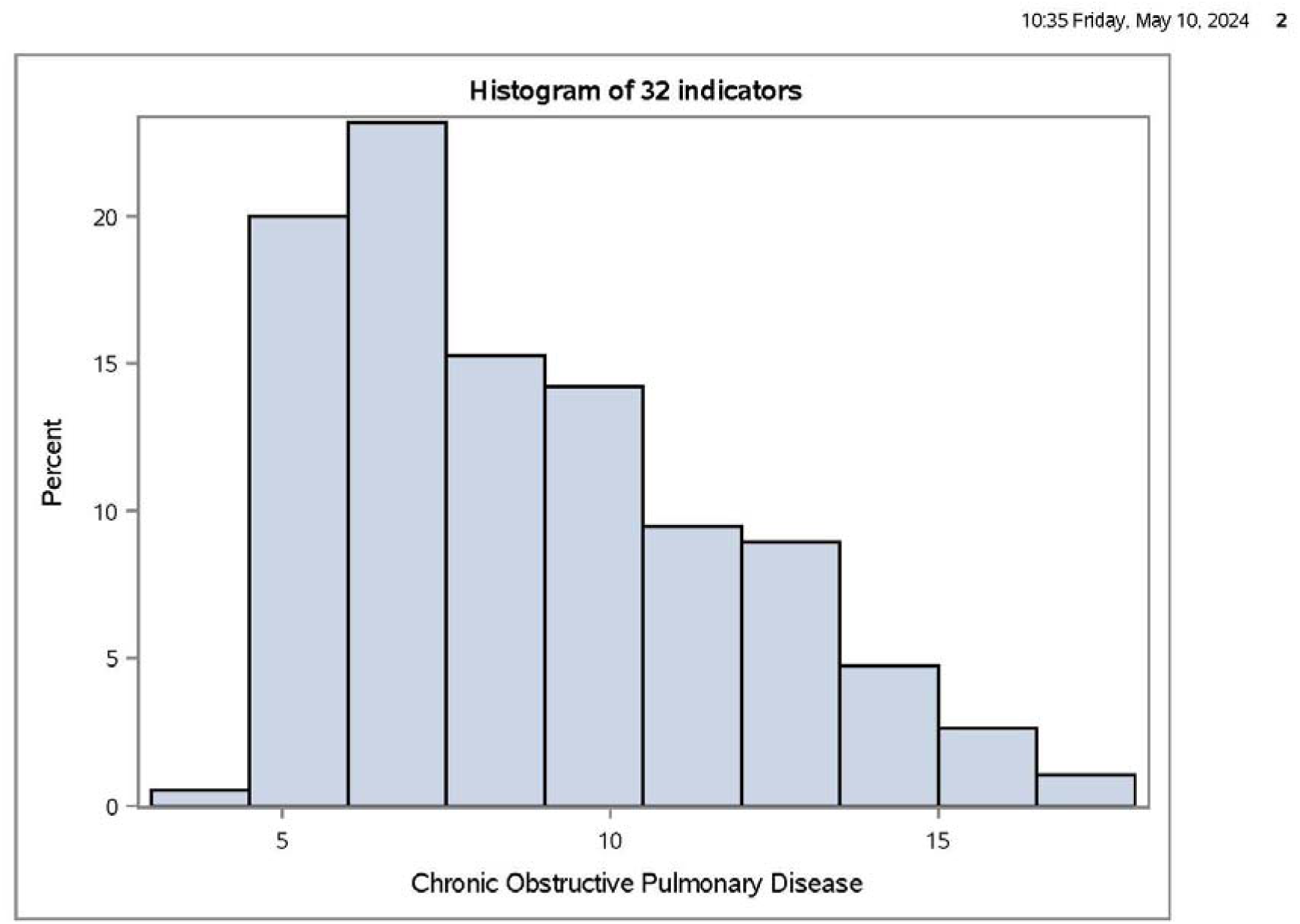
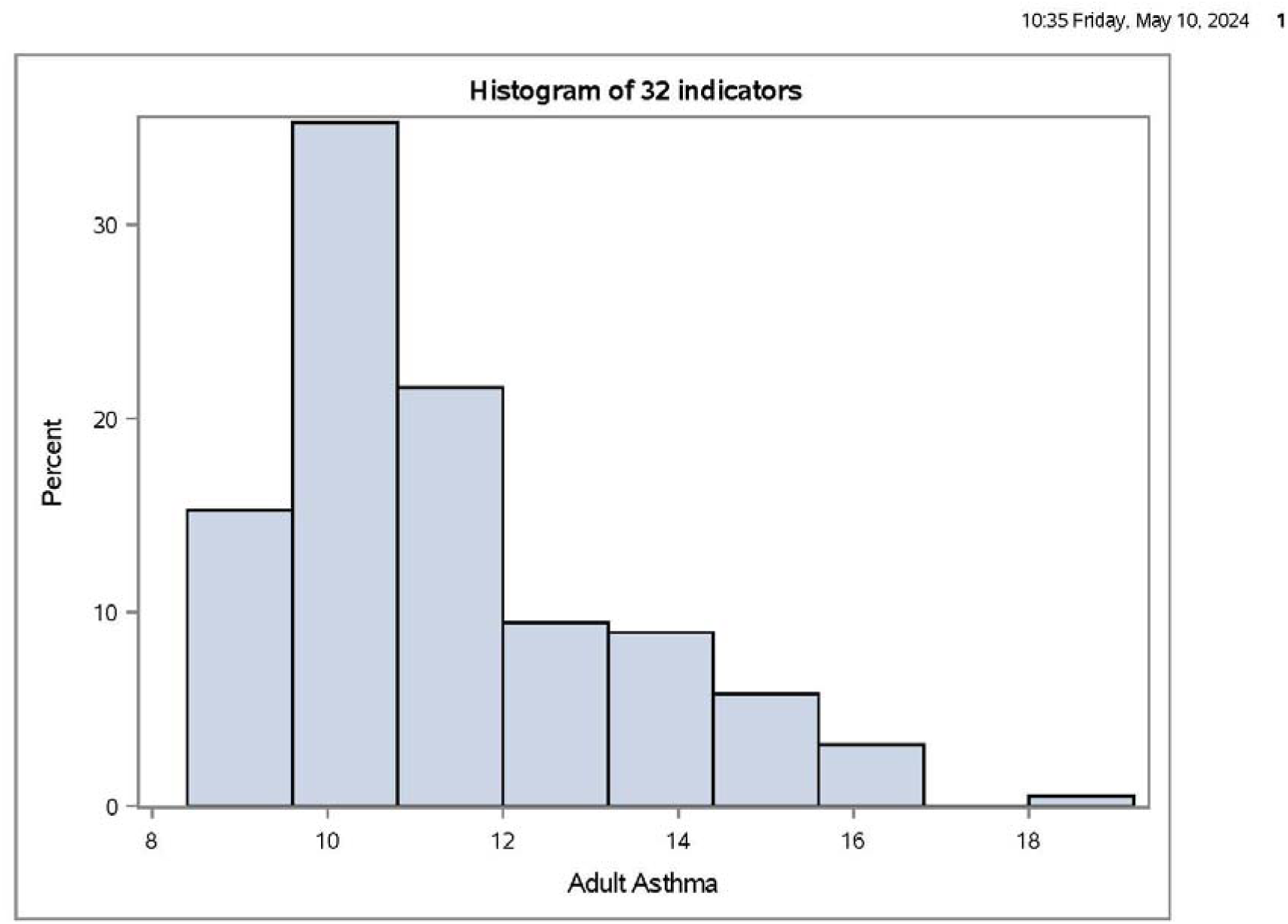
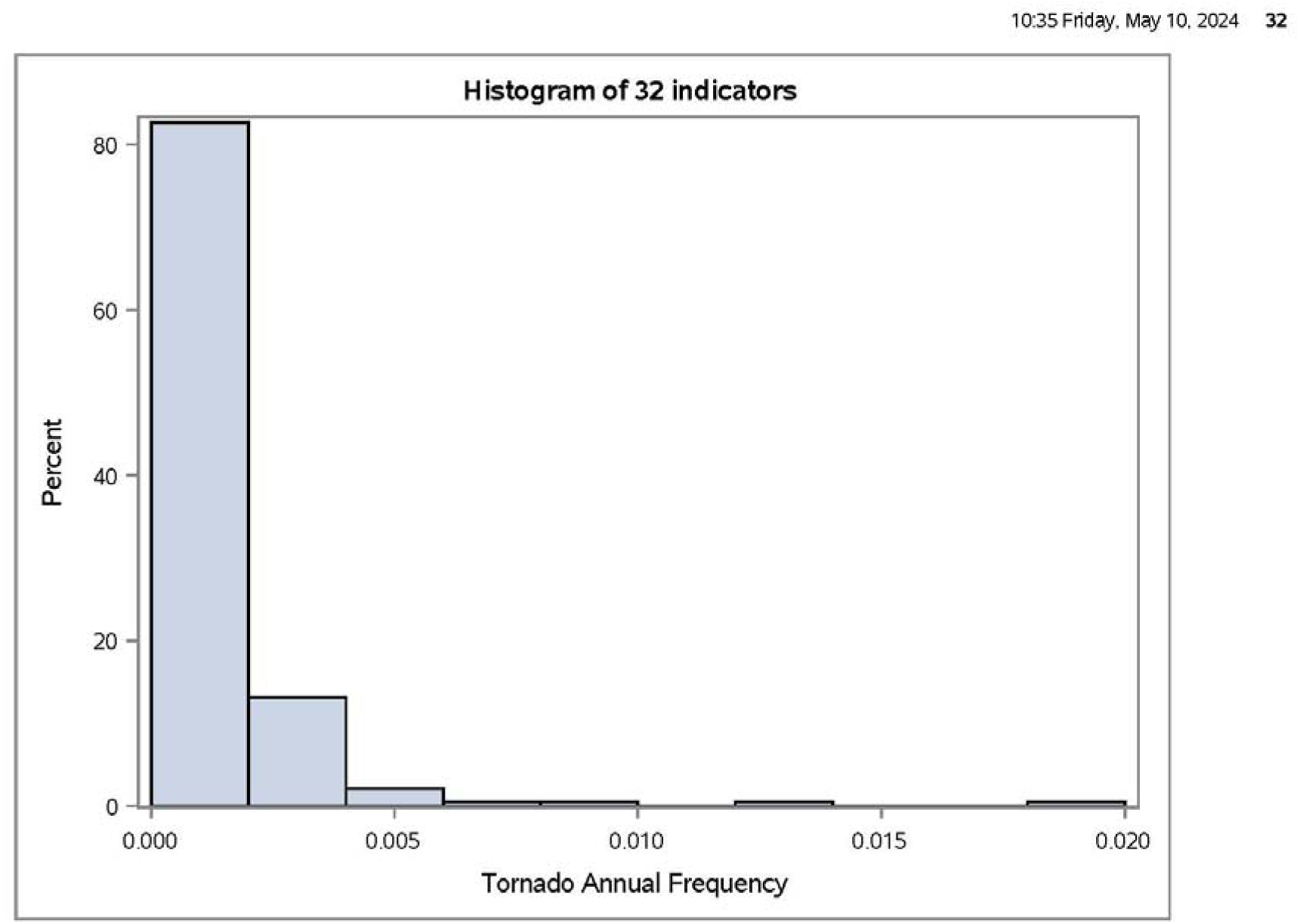
Heterogeneity analysis for the 32 indicators. Analysis was performed for each indicator to ensure only factors that offer significant variance were included in the index (32 panels).

## References

1. Bhandari S, Lewis PGT, Craft E, Marvel SW, Reif DM, Chiu WA. 2020. HGBEnviroScreen: enabling community action through data integration in the Houston– Galveston–Brazoria region. Int J Env Res Pub He 17(4):1130. 10.3390/ijerph17041130

2. Lewis PG, Chiu WA, Nasser E, Proville J, Barone A, Danforth C, et al. 2023. Characterizing vulnerabilities to climate change across the United States. Environ Int 172:107772. 10.1016/j.envint.2023.107772

3. Marvel SW, To K, Grimm FA, Wright FA, Rusyn I, Reif DM. 2018. ToxPi graphical user interface 2.0: Dynamic exploration, visualization, and sharing of integrated data models. BMC Bioinform 19:80. 10.1186/s12859-018-2089-2

4. Reif DM, Sypa M, Lock EF, Wright FA, Wilson A, Cathey T, et al. 2013. ToxPi GUI: an interactive visualization tool for transparent integration of data from diverse sources of evidence. Bioinformatics 29(3):402–3. 10.1093/bioinformatics/bts686

5. Glassman B. 2020. The multidimensional deprivation index using different neighborhood quality definitions. United States Census Bureau Social, Economic, and Housing Statistics Division. https://www.census.gov/content/dam/Census/library/working-papers/2020/demo/SEHSD-WP2020-08.pdf [accessed 24 April 2024].

6. Centers for Disease Control and Prevention. 2024. National Notifiable Diseases Surveillance System (NNDSS). https://www.cdc.gov/nndss/index.html [accessed 24 April 2024].

7. Casey JA, Su JG, Henneman LR, Zigler C, Neophytou AM, Catalano R, et al. 2020. Improved asthma outcomes observed in the vicinity of coal power plant retirement, retrofit and conversion to natural gas. Nature Energy 5(5):398–408. 10.1038/s41560-020-0600-2

8. Hilmers A, Hilmers DC, Dave J. 2012. Neighborhood disparities in access to healthy foods and their effects on environmental justice. Am J Public Health 102(9):1644–54. 10.2105/AJPH.2012.300865

9. Gochfeld M, Burger J. 2011. Disproportionate exposures in environmental justice and other populations: the importance of outliers. Am J Public Health 101(S1):S53–63. 10.2105/AJPH.2011.300121

10. Johnson DP, Stanforth A, Lulla V, Luber G. 2012. Developing an applied extreme heat vulnerability index utilizing socioeconomic and environmental data. Appl Geogr 35(1–2):23–31. 10.1016/j.apgeog.2012.04.006

11. Marvel SW, House JS, Wheeler M, Song K, Zhou YH, Wright FA, et al. 2021. The COVID-19 Pandemic Vulnerability Index (PVI) Dashboard: monitoring county-level vulnerability using visualization, statistical modeling, and machine learning. Environ Health Perspect 29(1):017701. 10.1289/EHP8690

12. Nasiri H, Yusof MJM, Ali TAM, Hussein MKB. 2019. District flood vulnerability index: urban decision-making tool. Int J Environ Sci Te 16(5):2249–58. 10.1007/s13762-018-1797-5

13. Newman G, Malecha M, Atoba K. 2023. Integrating ToxPi outputs with ArcGIS Dashboards to identify neighborhood threat levels of contaminant transferal during flood events. J Spat 68(1):57–69. 10.1080/14498596.2021.1891149

14. Xue J, Zartarian V, Tornero-Velez R, Stanek LW, Poulakos A, Walts A, et al. 2022. A generalizable evaluated approach, applying advanced geospatial statistical methods, to identify high lead exposure locations at census tract scale: Michigan case study. Environ Health Persp 130(7):077004. 10.1289/EHP9705

15. World Health Organization. 2000. Considerations in evaluating the cost-effectiveness of environmental health interventions. https://www.who.int/publications/i/item/WHO-SDE-WSH-00.10 [accessed 9 April 2024].

16. Bouzid M, Hooper L, Hunter PR. 2013. The effectiveness of public health interventions to reduce the health impact of climate change: a systematic review of systematic reviews. PLoS One 8(4):e62041. 10.1371/journal.pone.0062041

17. Environmental Defense Fund & Texas A&M University. 2023. The U.S. Climate Vulnerability Index. https://climatevulnerabilityindex.org/ [accessed 21 May 2024].

18. United States Census. Jefferson County, Kentucky. Population, Census, April 1, 2020. https://www.census.gov/quickfacts/fact/table/jeffersoncountykentucky/POP010220 [accessed 24 April 2024].

19. University of Kentucky. 2023. Kentucky Annual Economic Report 2023. https://cber.uky.edu/sites/cber/files/publications/UK%20CBER%20Kentucky%20Annual%20Economic%20Report%202023_Web.pdf [accessed 3 May 2024].

20. United States Climate Resilience Toolkit. The Climate and Economic Justice Screening Tool (CEJST) https://toolkit.climate.gov/tool/climate-and-economic-justice-screening-tool [accessed 9 April 2024].

21. Centers for Disease Control and Prevention. 2021. Places: Census Tract Data – GIS Friendly Format https://chronicdata.cdc.gov/500-Cities-Places/PLACES-Census-Tract-Data-GIS-Friendly-Format-2021-/yjkw-uj5s/data [accessed 1 Nov 2023].

22. Kentucky Cabinet for Health and Family Services. Division of Health Care Facilities. https://chfs.ky.gov/agencies/os/oig/dhc/Pages/hcf.aspx [accessed 2 November 2023].

23. National Center for Health Statistics. 2018. U.S. Small-Area Life Expectancy Estimates Project (USALEEP): Life Expectancy Estimates File for Kentucky, 2010-2015. https://www.cdc.gov/nchs/nvss/usaleep/usaleep.html [accessed 1 November 2023].

24. United States Department of Housing and Urban Development. 2023. Environmental Health Hazard Index. https://data.lojic.org/datasets/HUD::environmental-health-hazard-index/about [accessed 9 November 2023].

25. United States Environmental Protection Agency. 2020. Chemical concentrations, exposures, health risks by census tract from National Scale Air Toxics Assessment (NATA). https://catalog.data.gov/dataset/chemical-concentrations-exposures-health-risks-by-census-tract-from-national-scale-air-toxics [accessed 9 November 2023].

26. United States Department of Transportation. 2020. National Transportation Noise Map. https://www.bts.gov/geospatial/national-transportation-noise-map [accessed 2 November 2023].

27. United States Environmental Protection Agency. 2021. Daily Census Tract-Level PM2.5 Concentrations, 2016 - 2020 https://data.cdc.gov/Environmental-Health-Toxicology/Daily-Census-Tract-Level-PM2-5-Concentrations-2016/96sd-hxdt/about_data [accessed 8 November 2023].

28. United States Environmental Protection Agency. 2021. Risk-Screening Environmental Indicators (RSEI) Model RSEI Version 2.3.11 (RY 2021). https://edap.epa.gov/public/extensions/EasyRSEI/EasyRSEI.html [accessed 2 November 2023].

29. United States Environmental Protection Agency. 2023. Facility Registry Service Facilities State Single File CSV Download. https://www.epa.gov/frs/epa-frs-facilities-state-single-file-csv-download [accessed 16 November 2023].

30. Kentucky Energy and Environmental Cabinet. Superfund Site List. https://dep.gateway.ky.gov/eSearch/AvailableReports/Details?id=47 [accessed 14 November 2023].

31. United States Environmental Protection Agency. 2021a. EJScreen: Environmental Justice Screening and Mapping Tool. Traffic proximity and volume dataset. https://gaftp.epa.gov/EJScreen/ [accessed 16 November 2023].

32. United States Geological Survey, U.S. Department of the Interior, and U.S. Forest Service. 2021. Tree Canopy Cover of the Conterminous United States, 2021 NLCD 2021 USFS Tree Canopy Cover (CONUS). Multi-Resolution Land Characteristics (MRLC) Consortium. https://www.mrlc.gov/data/nlcd-2021-usfs-tree-canopy-cover-conus [accessed 11 November 2023].

33. United States Department of Housing and Urban Development. 2023. Location Affordability Index (Version 3). https://data.lojic.org/datasets/HUD::location-affordability-index-v-3/about [accessed 17 November 2023].

34. Centers for Disease Control and Prevention. 2020. Agency for Toxic Substances and Disease Registry/ Geospatial Research, Analysis, and Services Program. CDC/ATSDR Social Vulnerability Index, Database Kentucky. https://www.atsdr.cdc.gov/placeandhealth/svi/data_documentation_download.html. Accessed on 20 November 2023.

35. United States Department of Agriculture, Economic Research Service. 2019. Food Access Research Atlas. https://www.ers.usda.gov/data-products/food-access-research-atlas/download-the-data/ [accessed 17 November 2023].

36. Esri. 2020. USA_Flood_Hazard_Reduced_Set_gdb. ArcGIS Online Map Viewer. https://www.arcgis.com/apps/mapviewer/index.html?layers=bf9585afc2934b648f3918693285b7c8 [accessed 20 November 2023].

37. Jefferson County Information Consortium. 2022. Jefferson County Kentucky Urban Heat Management Study (LOJIC). https://hub.arcgis.com/datasets/LOJIC::jefferson-county-ky-urban-heat-management-study [accessed 8 May 2024].

38. Federal Emergency Management Agency. 2023. National Risk Index (NRI) Data & Resources. https://hazards.fema.gov/nri/data-resources [accessed 23 September 2023].

39. Toxicological Prioritization Index (ToxPi) version 1.2.1 (https://CRAN.R-project.org/package=toxpiR)

40. Wang R, Browning MH, Kee F, Hunter RF. 2023. Exploring mechanistic pathways linking urban green and blue space to mental wellbeing before and after urban regeneration of a greenway: Evidence from the Connswater Community Greenway, Belfast, UK. Landscape Urban Plan 235:104739. 10.1016/j.landurbplan.2023.104739

41. O’Cathain A, Croot L, Duncan E, Rousseau N, Sworn K, Turner KM, et al. 2019. Guidance on how to develop complex interventions to improve health and healthcare. BMJ Open 9(8):e029954. 10.1136/bmjopen-2019-02995

42. Louisville Metro-Jefferson County Government. Louisville Urban Tree Canopy Assessment, 2015. https://louisvilleky.gov/urban-forestry/document/louisville-urban-tree-canopy-assessment-2015 [accessed 9 April 2024].

43. Messer LC, Laraia BA, Kaufman JS, Eyster J, Holzman C, Culhane J, et al. 2006. The development of a standardized neighborhood deprivation index. J Urban Health 83(6):1041–62. 10.1007/s11524-006-9094-x

44. Brown AL, Van Kamp I. 2017. WHO environmental noise guidelines for the European Region: A systematic review of transport noise interventions and their impacts on health. Int J Env Res Pub He 14(8):873. 10.3390/ijerph14080873

45. Hammer MS, Swinburn TK, Neitzel RL. 2014. Environmental noise pollution in the United States: developing an effective public health response. Environ Health Perspect 122(2):115–9. 10.1289/ehp.1307272.

46. Uong SP, Zhou J, Lovinsky-Desir S, Albrecht SS, Azan A, Chambers EC, et al. 2023. The creation of a multidomain neighborhood environmental vulnerability index across New York City. J Urban Health 100(5):1007–23. 10.1007/s11524-023-00766-3

47. Resnik DB, Zeldin DC, Sharp RR. 2005. Research on environmental health interventions: ethical problems and solutions. Accountability Res 12(2):69–101. 10.1080/08989620590957157

48. Anderson LB, Ness HD, Holm RH, Smith T. 2024. Wastewater-informed digital advertising as a Covid-19 geotargeted neighborhood intervention: Jefferson County, Kentucky, 2021–2022. Am J Public Health 114(1):34–7. 10.2105/AJPH.2023.307439

49. Ho HC, Wong MS, Man HY, Shi Y, Abbas S. 2019. Neighborhood-based subjective environmental vulnerability index for community health assessment: development, validation and evaluation. Sci Total Environ 654:1082–90. 10.1016/j.scitotenv.2018.11.136

